# Multimodal Hyperbolic Graph Learning for Alzheimer’s Disease Detection

**DOI:** 10.1101/2024.10.29.24316334

**Authors:** Chengyao Xie, Wenhao Zhou, Ciyuan Peng, Azadeh Noori Hoshyar, Chengpei Xu, Usman Naseem, Feng Xia

## Abstract

Multimodal graph learning techniques have demonstrated significant potential in modeling brain networks for Alzheimer’s disease (AD) detection. However, most existing methods rely on Euclidean space representations and overlook the scale-free and small-world properties of brain networks, which are characterized by power-law distributions and dense local clustering of nodes. This oversight results in distortions when representing these complex structures. To address this issue, we propose a novel multimodal Poincaré Fréchet mean graph convolutional network (MochaGCN) for AD detection. MochaGCN leverages the exponential growth characteristics of hyperbolic space to capture the scale-free and small-world properties of multimodal brain networks. Specifically, we combine hyperbolic graph convolution and Poincaré Fréchet mean to extract features from multimodal brain networks, enhancing their representations in hyperbolic space. Our approach constructs multimodal brain networks by integrating information from diffusion tensor imaging (DTI) and functional magnetic resonance imaging (fMRI) data. Experiments on the Alzheimer’s Disease Neuroimaging Initiative (ADNI) dataset demonstrate that the proposed method outperforms state-of-the-art techniques.

## 1 Introduction

Alzheimer’s disease (AD) is an irreversible neurodegenerative disorder that impacts cognition, function, and behavior, progressively leading to a loss of physical functions and, ultimately, death [14]. Approximately 60 million people world-wide were affected by Alzheimer’s disease [15], and the global economic burden is projected to reach 9.12 trillion US dollars by 2050 [9]. These staggering figures underscore the urgent need for more effective diagnostic and monitoring tools. The disease is linked with the accumulation of amyloid beta and tau proteins in specific cortex areas, causing the loss of neurons and synapses. Neuroimaging that reflects the structural, functional connectivity properties of the human brain can be used to analyze disease development.

Recently, graph learning techniques have demonstrated significant capabilities in brain network modeling, achieving promising results in brain disorder detection [5, 17, 19, 22, 23]. With the increasing availability of multimodal neuroimaging data, such as functional magnetic resonance imaging (fMRI), diffusion tensor imaging (DTI), computed tomography (CT), and positron emission tomography (PET), multimodal brain graph learning has become a trending research area. Some works enhance the detection performance by integrating graph neural network (GNN) and its variants with novel multimodal feature learning [11, 25]. In addition, some studies highlight that the graph structure information is crucial for advancing the brain disorder detection performance [4, 12, 27]. For example, Zhang et al. [29] proposed a multimodal graph neural network to fuse the learned node representations for AD prediction. However, these methods rely on Euclidean space representations, which are not suitable for capturing the scale-free and small-world properties of brain networks [26, 30], as shown in Fig. 1. As brain regions are organized hierarchically, the regions and their connections grow exponentially. This exponential growth surpasses the polynomial expansion capacity of Euclidean space, leading to **representation distortion**.

**Fig. 1:**
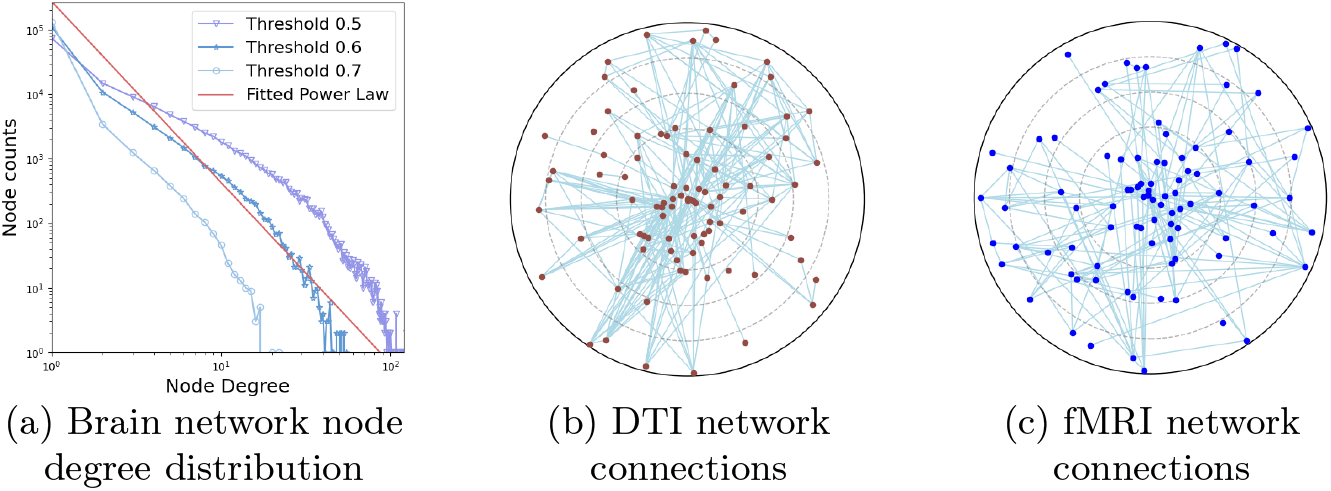
Scale-free and small-world properties of brain networks. (a) The node degree distribution of the fMRI brain networks from 115 subjects in the ADNI dataset follows a power-law distribution, indicates the scale-free characteristics. The visualizations of (b) the DTI network and (c) the fMRI network projected onto the Poincaré disk for one subject demonstrate small-world characteristics.

Notably, hyperbolic geometry can describe data structures that grow exponentially more effectively than Euclidean space [16, 30]. Recent studies use hyperbolic graph learning to achieve superior representations and improve model availability [1, 18, 20, 28]. For example, Zhang et al. [28], projected the functional and structural brain network into hyperbolic space and utilized a hyperbolic graph kernel for the diagnosis of mild cognitive impairment (MCI). These works demonstrate that hyperbolic geometry can better capture graph structures and improve model performance.

Therefore, to reduce representation distortion of multimodal brain networks and enhance information fusion, we propose a novel multimodal hyperbolic graph learning method (MochaGCN) for AD detection. Specifically, we introduce a hyperbolic graph convolutional network with Poincaré Fréchet mean aggregation to capture multimodal brain network features in hyperbolic space. We fuse information from DTI and fMRI modalities and employ contrastive learning to enhance the fused features by minimizing the contrastive loss. Finally, we calculate the hyperbolic distance of the learned representation features for AD detection. The proposed model offers a more accurate representation of multimodal brain networks, holding significant implications for the diagnosis and treatment of brain disorders.

Our contributions can be summarized as follows:

– We propose a novel multimodal hyperbolic Poincaré Fréchet mean graph convolutional neural network (MochaGCN) for AD detection, significantly reducing the representation distortion of brain networks and enhancing information fusion between DTI and fMRI modalities.
– We combine a graph convolutional network with the Poincaré Fréchet mean aggregation to preserve scale-free and small-world properties of multimodal brain network representations in hyperbolic space.
– The experimental results on the Alzheimer’s Disease Neuroimaging Initiative (ADNI) dataset demonstrate the effectiveness of the proposed method, which outperforms the state-of-the-art methods.

## 2 Preliminaries

### 2.1 Problem Definition

The AD detection task first obtains the connectivity from preprocessed DTI and fMRI neuroimages to construct the brain networks. The networks are represented as graph *𝒢*_DTI_ = (*V*_DTI_, *E*_DTI_) and 𝒢 _fMRI_ = (*V*_fMRI_, *E*_fMRI_), where *V* is the set of vertices representing brain regions and *E* is the set of edges connecting the regions. The connectivity graph in Euclidean space is represented by an adjacency matrix 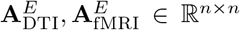, where *n* is the number of regions of interest (ROIs). Each element *e*_*i,j*_ of **A** represents the connectivity strength between regions *i* and *j*. Each modality’s graphs form the data **M**_DTI_ and **M**_fMRI_, where **M** = *{***A**_1_, **A**_2_, …, **A**_*n*_*p }* and *n*_*p*_ is the number of participants. The goal of this research is to learn a classification function 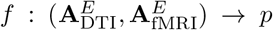, where *p* is the probability of subject class *γ*, i.e., *γ* = 1 for AD patients and *γ* = 0 for normal controls. The function *f* (*·*) is learned from a training set of subjects with known labels. The learned function is then used to predict the labels of new subjects in a test set and its performance is evaluated using standard classification metrics.

### 2.2 Hyperbolic Geometry

Hyperbolic geometry, characterized by negative curvature, differs from the flat Euclidean space. Hyperbolic space grows exponentially. Specifically, considering a 2-dimensional hyperbolic disk with radius *r* and constant curvature -1, the perimeter and area of the disk can be calculated by 2*π* sinh *r* and 2*π*(cosh *r−* 1), and both of them grow as *e*^*r*^ with *r*, compared with the linearly and quadratically growth in Euclidean space. So it is ideal for representing data with exponential expansion and hierarchical structures. There are many models to embed hyperbolic space in Euclidean space. And in this paper, the Poincaré model is introduced as the necessary background of this study. Hyperbolic manifold ℍ with curvature *K <* 0 can be mapped to Euclidean space by the Poincaré ball model, which is defined as:

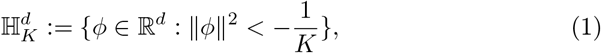

where ∥ · ∥ denotes the Euclidean norm. When lim *K* = 0, the hyperbolic space is isometric to the Euclidean space. For two points *ϕ*_1_, *ϕ*_2_ in the hyperbolic space, the Möbius addition is defined as:

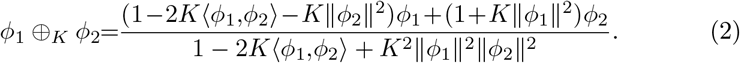

And the distance between them is defined as

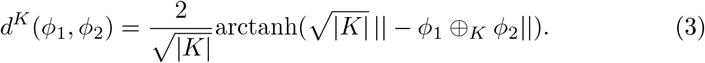

For a point *ϕ* in the hyperbolic space, the tangent space 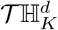 at *ϕ* is Euclidean space. The coordinates can be mapped to each other by the exponential map and logarithmic map. The exponential map *expK* : 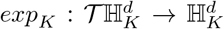 maps a vector *ϕ*^*E*^ in the tangent space to a point in the hyperbolic space, and the logarithmic map 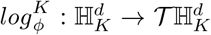 maps a point *ϕ*^*H*^ in the hyperbolic space to a vector in the tangent space. For a given point *ϕ*, the exponential map of point *ϕ*^*E*^ and logarithmic map of point *ϕ*^*H*^ are defined as:

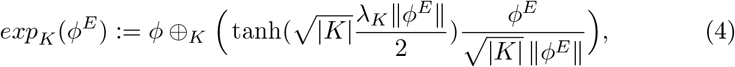

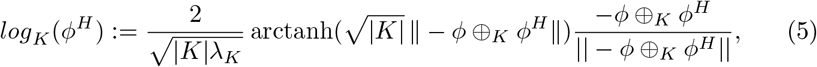

where *λ*_*K*_ = 2(1 + K ∥ *ϕ* ∥^2^)^*−*1^ is the conformal factor.

According to [6], we formulate matrix-vector multiplication, bias translation, and activation function of hyperbolic neural networks are defined as:

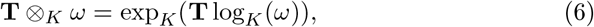

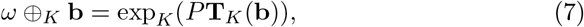

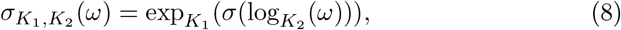

respectively, where **T** is a matrix, *ω* is a vector, **b** is a bias, and *σ* is an activation function.

## 3 Methodology

The AD detection task is a binary classification problem and involves four main steps. First, functional, structural, and fused brain networks are constructed for each subject based on DTI and fMRI imaging data. Next, hyperbolic graph convolution is applied to these brain networks to extract representation feature vectors. Then, contrastive learning minimizes the contrastive loss between the fused vector and the functional and structural vectors. Finally, the fused feature vectors are projected into hyperbolic space, and the distance from the cluster center is calculated to generate the diagnosis results. The overall architecture of the proposed method is shown in Fig. 2.

**Fig. 2:**
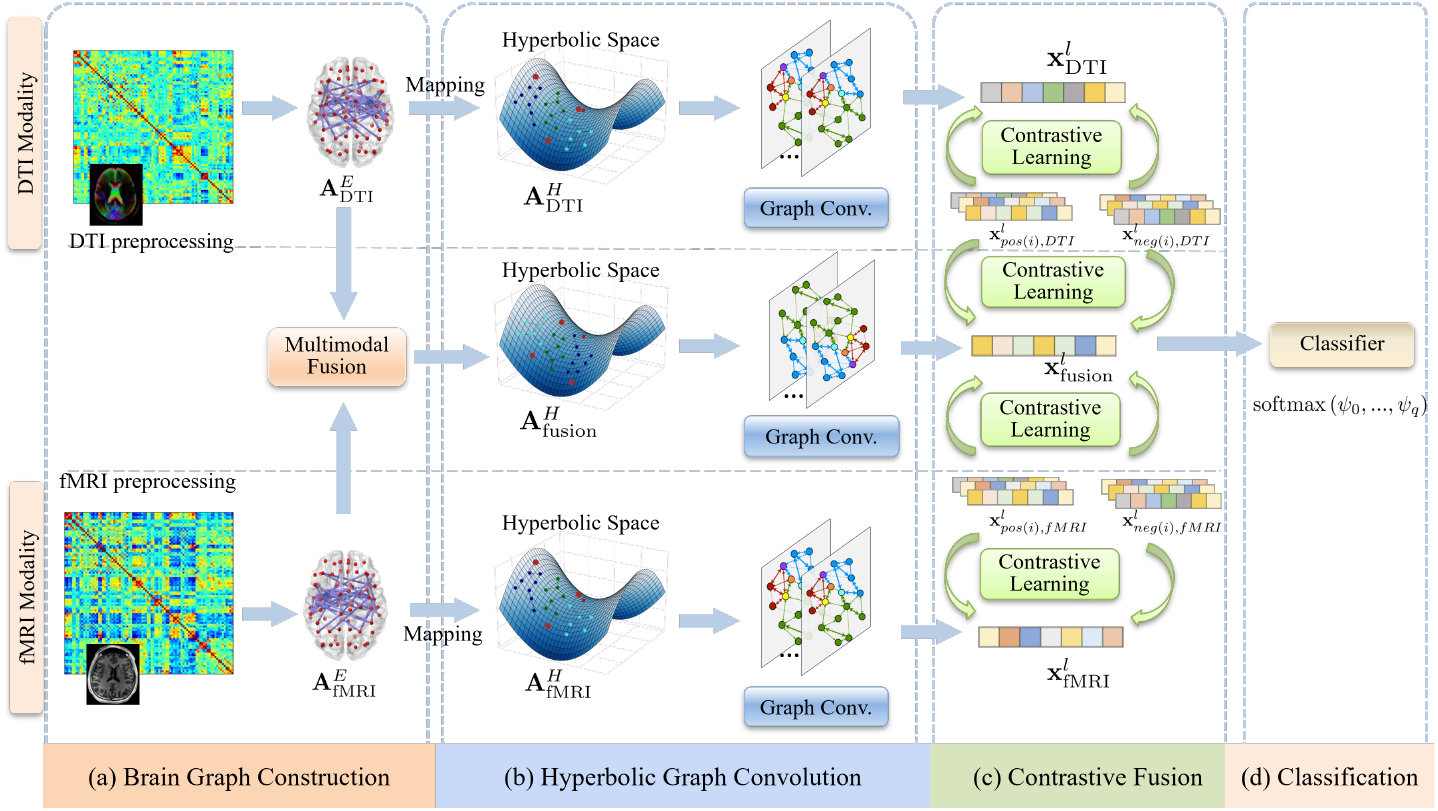
Overall framework of MochaGCN. (a) Calculation of the fMRI functional connectivity matrix and DTI structural connectivity matrix based on the AAL brain atlas, followed by graph construction. (b) Projection of the brain graph to hyperbolic space and fusion of the DTI fMRI information into a fusion modality, with hyperbolic Poincaré Frechet mean graph convolution (Graph Conv.) applied to each modality. (c) Feature learning for DTI and fMRI modalities via contrastive learning, where the fusion feature is learned by contrasting the fusion feature with the other two features. (d) Disease classification by calculating the fusion feature’s hyperbolic distance.

### 3.1 Brain Graph Construction

We begin by preprocessing the neuroimaging data to construct structural graph 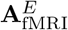 derived from DTI data, and functional graph 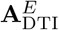 based on fMRI data. Both graphs are initially represented in Euclidean space. Subsequently, we employ the mapping techniques outlined in Section 3.2 to project these graphs into hyperbolic space, and get hyperbolic counterparts 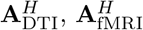.

We introduce a multimodal fusion graph to leverage both early and late fusion strategies [13]. In the early fusion stage, we integrate the information from both modalities in hyperbolic space. This fusion is formulated as follows:

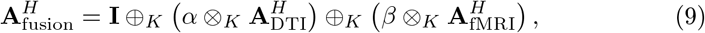

where **I** is the identity matrix. *α, β* are the fusion parameters, and *α* + *β* = 1.

### 3.2 Hyperbolic Graph Convolution

To capture the brain graph features for AD detection, a hyperbolic graph convolutional network will be used to learn the representation of the brain network. The input graphs are represented by a binary adjacency matrix and the outputs are hyperbolic embeddings for graph classification and link prediction tasks. The proposed model architecture is illustrated in Fig. 2. The model consists of the following parts:

– **Mapping** We first transport the input data between Euclidean space and hyperbolic space via logarithmic map Eq. (5) and exponential map Eq. (4). For DTI and fMRI brain networks, we first map the input data to hyperbolic space using the exponential map 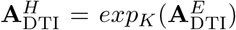 and 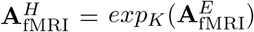, where **A**^*H*^ represents the brain graph in hyperbolic space.
– **Node Aggregation** Aggregation function captures features of neighborhood in the graph, and is a core component of convolutional networks. Lou et al. [7] extended Fréchet mean for arbitrary Riemannian manifolds, and Cao [2] presented a (1− *ϵ*)-approximation to fix imprecise convergences problem. In this work, we combine the Poincaré Fréchet mean and hyperbolic graph convolution to aggregate the information from neighboring nodes. Each modality adjacent matrix in hyperbolic space, 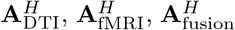 is then fed into the hyperbolic graph convolution module to learn the corresponding representations. This process is formulated as:

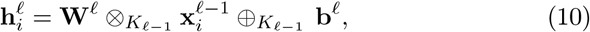

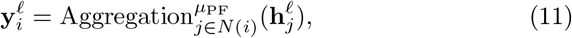

where *ℓ* is the layer index, where 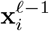 are the features of node *i* of the previous layer *ℓ −* 1, and **W**^*ℓ*^ and **b**^*ℓ*^ the weights and bias parameters of layer *ℓ, N* (*i*) denotes the neighbors of node *i*. The Poincaré Fréchet mean *µ*_PF_ is defined as:

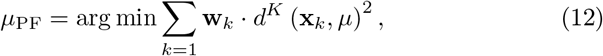

where *d*^*K*^ is the the hyperbolic distance as introduced in Eq. (3), *w*_*k*_ is the weight of the *k*-th point, and *µ* is the Fréchet mean of the points in the manifold.
– **Activation Function** We implement the hyperbolic ReLU function using Eq. (8). The output was defined by:

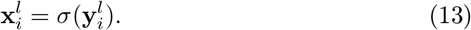

The output of each modality is 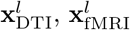, and 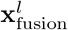 respectively, where *l* is the number of layers.

### 3.3 Contrastive Fusion

According to [13], we will leverage late contrastive fusion techniques, further enhancing the fusion modality features based on previously learned representations 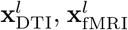, and 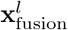. For the DTI modality, we select subjects with the same class labels as positive samples and those with different class labels as negative samples from the dataset and follow the approach of SimCLR [3] and HCL [24] to construct a contrastive loss, as follows:

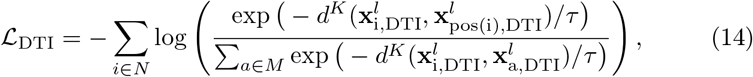

where *τ ∈ R*^+^ is a scalar temperature parameter, *M* denotes a batch samples, **x**_*pos*(*i*)_ is the positive pairs of node *i*. We can also obtain a contrastive loss *ℒ* _fMRI_ for fMRI modality in a similar way. These two contrastive losses are used to update the DTI and fMRI representations separately without exchanging information with each other.

Since the fused modality integrated the information from both modalities early on, it will further refine this information by comparing to the features extracted from DTI and fMRI modalities in the late stage. The fusion modality contrastive loss can be calculated by:

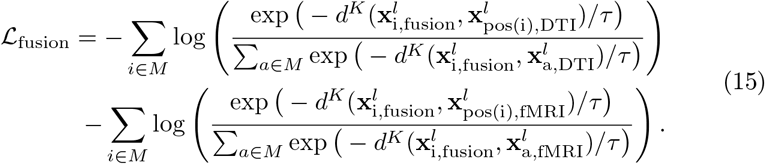

#### Algorithm 1

Learning process of MochaGCN

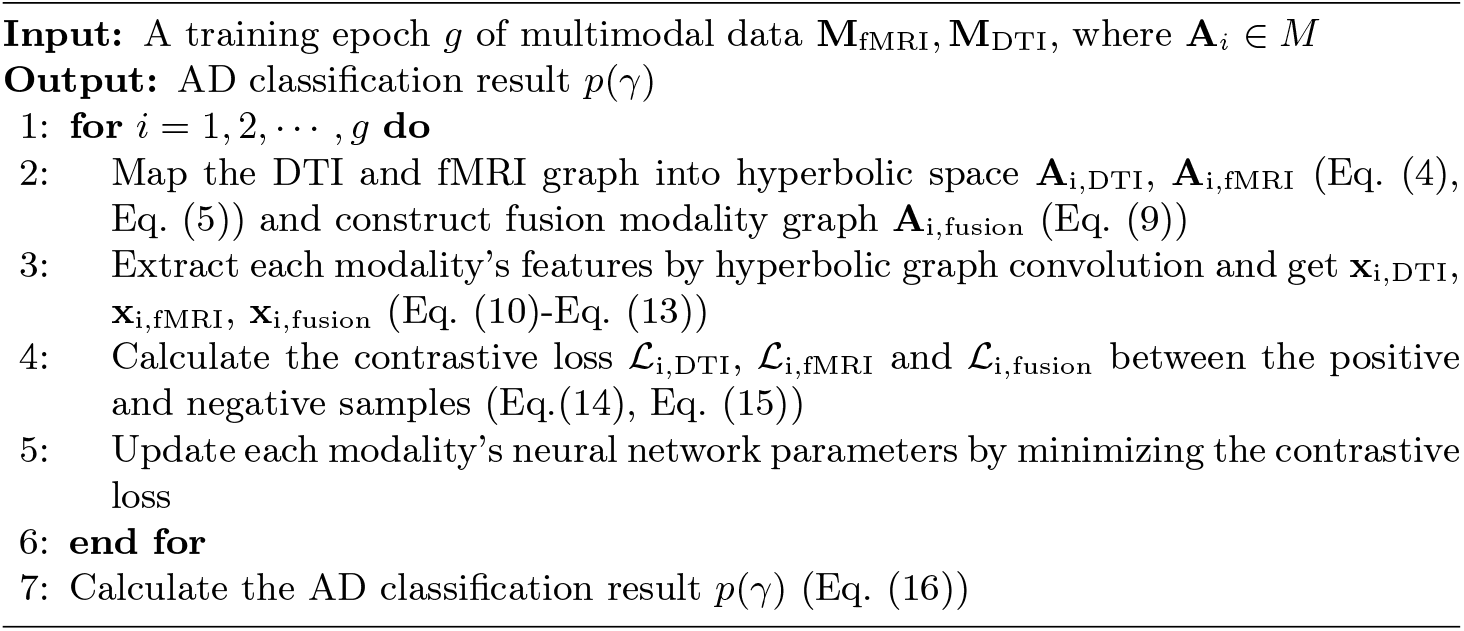

### 3.4 Classification

The AD classification probability is calculated using the hyperbolic distance between the fusion modality’s extracted features and the centers *𝒞* = [**c**_1_, **c**_2_, …, **c**_*q*_] of different classes, where *q* is the number of classes. The probability of the *j*-th class is calculated as follows:

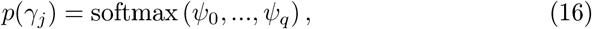

where *ψ*_*i*_ = (exp(*d*^*K*^(**x**_fusion_, **c**_*i*_)^2^ *−* 2) + 1)^*−*1^ and *γ*_*j*_ is the *j*-th class.

To elucidate the data processing mechanism of MochaGCN, we employ pseudocode to describe the model’s AD classification process, as outlined in Algorithm 1.

### 3.5 Complexity Analysis

The time complexity of the primary component of MochaGCN is analyzed below. The complexity for mapping the brain graph to hyperbolic and tangent spaces is *O*(*n*^2^), where *n* represents the number of ROIs. The complexity for forward training and backpropagation in the Poincaré Fréchet mean graph convolution is *O*(*L·* (*n· f* ^2^+*T· n· d*)), where *L* denotes the number of layers, *f* is the dimension of embedding features, *T* is number of iterations for Fréchet mean convergence, and *d* is average degree of the nodes. The complexity of the contrastive fusion module is *O*(*n · f*). The remainder no longer involves high computational complexity.

## 4 Experiments

### 4.1 Dataset and Experimental Settings

The Alzheimer’s Disease Neuroimaging Initiative (ADNI) ^6^ dataset is a comprehensive neuroimaging dataset for AD diagnosis. We evaluate 115 subjects, with 60 normal controls (NC), and 55 patients with AD for the experiments. Each participant has both DTI and fMRI data.

To ensure the reproducibility of this study, we adopt QSIprep 0.19.1 and fMRIprep 20.2.3 to preprocess DTI and fMRI modalities respectively. We analyze brain networks using the Automated Anatomical Labeling (AAL) Atlas, which contains 90 ROIs. Then the Pearson correlation coefficient and fractional anisotropy are used for fMRI and DTI brain graph construction. The hyperbolic graph convolutional model used in this work has two layers, with the curvature set to -1.0 and an embedding dimension of 128. During training, we apply a learning rate of 0.01 and the Adam optimizer to update network weights. To evaluate the model’s performance, we conducted 5-fold cross-validation, repeating the experiments 10 times to ensure robust results.

### 4.2 Baselines

To establish a benchmark for comparison, we examined existing studies that leverage DTI and fMRI data for AD diagnosis. These studies incorporate advanced deep learning techniques, such as dynamic graph learning and hypergraph learning, to improve diagnostic accuracy. The state-of-the-art baseline methods are as follows:

– DecGAN [10] employs a decoupling generative adversarial network to detect abnormal neural circuits associated with AD.
– mmLasso [8] utilizes a multi-modal LassoNet framework to integrate supplementary information from different modalities.
– Cross-GNN [21] captures inter-modal dependencies through dynamic graph learning and mutual learning.
– PALH [31] predicts abnormal brain connections through a prior-guided adversarial learning framework with hypergraph.

### 4.3 Result Analysis

We evaluate the performance of the model using accuracy (ACC), sensitivity (Sen.), and specificity (Spe.). The mean values achieved are 93.56% for accuracy, 95.34% for sensitivity, and 91.54% for specificity, with standard deviations (SD) of 1.10%, 2.10%, and 3.64% respectively, as shown in Table 1. Since the baseline methods do not provide open-source code, their results were collected from their original papers. Our model achieved the highest accuracy and demonstrates strong overall classification performance and effectiveness. For example, while mmLasso [8] uses 16,830 dimensions for disease classification, our model only uses 128 dimensions, significantly reducing the feature space without compromising performance. Although our model’s sensitivity and specificity were slightly below the best, it maintains a strong balance between correctly identifying true positives (sensitivity) and minimizing false positives (specificity), ensuring reliable disease detection. Overall, MochaGCN outperforms the baseline models in detecting Alzheimer’s disease with improved accuracy and efficiency.

**Table 1:** Classification performance comparison with baseline methods.

| Method | ACC(%) | Sen.(%) | Spe.(%) |
| --- | --- | --- | --- |
| DecGAN [10] | 85.18 | 81.25 | 90.91 |
| mmLasso [8] | 90.68±0.34 | 88.81±0.68 | 91.91±0.55 |
| Cross-GNN [21] | 88.6±6.3 | **95.9**±9.2 | 85.2±7.7 |
| PALH [31] | 88.73 | 84.37 | **92.31** |
| MochaGCN | **93.56**±1.10 | 95.34±2.10 | 91.54±3.64 |

### 4.4 Effectiveness Analysis of Hyperbolic Representation

#### Effect of Hyperbolic Curvature

To evaluate the impact of hyperbolic curvature on the performance of our model, we conducted experiments across different curvature values, and curvature 0 means Euclidean space. This test is conducted with embedding dimension 16. The results are shown in Fig. 3(a). The hyperbolic and Euclidean results are colored blue and orange, respectively. Overall, our model with negative curvature outperforms the flat curvature in the classification task, suggesting that hyperbolic space representation is a superior choice for multimodal fusion in AD diagnosis.

**Fig. 3:**
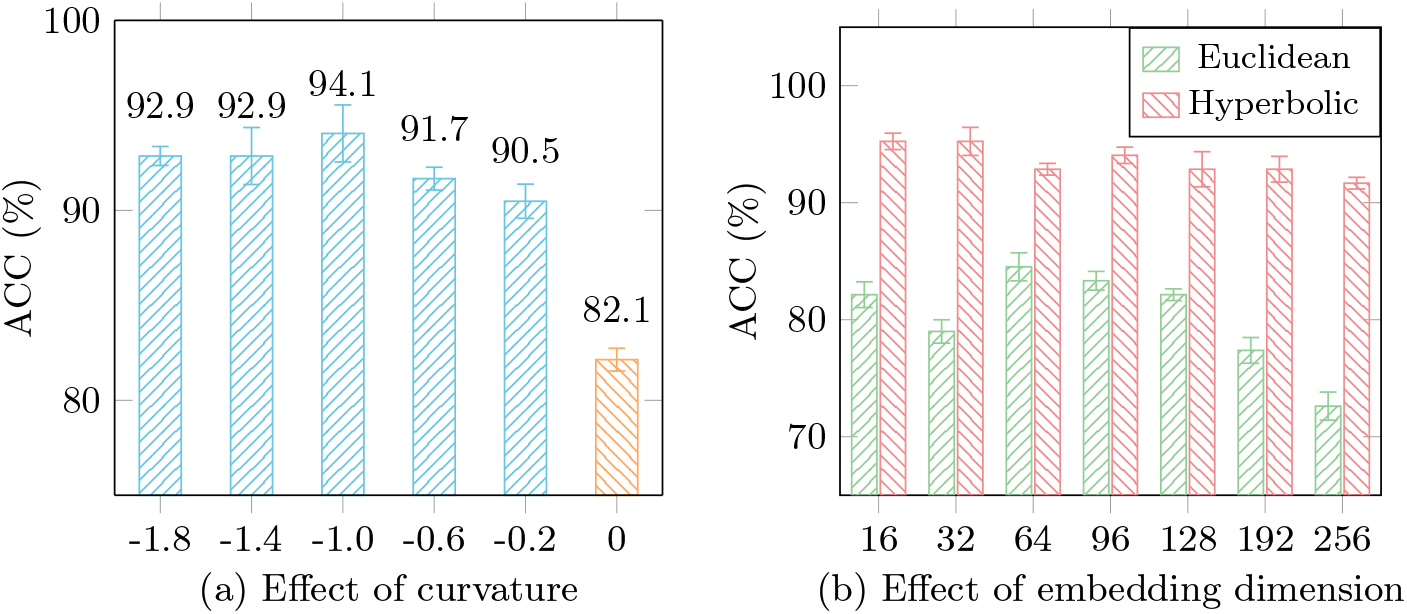
Performance comparison of different curvature values (a) and dimensions (b). In (a), the best result is observed at a curvature of -1.0, and worst is at curvature 0 which corresponds to Euclidean space. In (b), the ACC (%) of the algorithm in Euclidean and hyperbolic space is compared.

#### Effect of Embedding Dimension

We conducted experiments with different embedding dimensions and also reimplemented the MochaGCN using GCN in Euclidean space for comparison. The hyperbolic curvature is set to -0.8. The results are shown in Fig. 3(b). Our proposed algorithm achieves near-optimal performance with an embedding dimension of 16, which is significantly smaller than the 64-dimensional embedding required by GCN. This indicates that hyperbolic space embedding can significantly reduce the required embedding dimension while maintaining high performance.

### 4.5 Ablation Study

To evaluate the contributions of each component in the proposed model, we conducted an ablation study by sequentially replacing the hyperbolic space convolution with the Euclidean space convolution, removing the fusion modality, and both. The hyperbolic curvature is set to -1.0 and embedding dimension is 128. The accuracy decreased by 11.42%, 4.99%, and 17.77%, respectively, as shown in Table 2. These results indicate that hyperbolic Poincaré Fréchet mean convolution significantly contributes to the overall performance of the model, also means proposed MochaGCN can greatly reduce the representation distortion compared to Euclidean space.

**Table 2:** Contribution of each component.

| Method | ACC(%) | Sen.(%) | Spe.(%) |
| --- | --- | --- | --- |
| MochaGCN | 93.56 | 95.34 | 91.54 |
| MochaGCN<sub>wo:hyperbolic</sub> | 82.14 | 81.11 | 81.79 |
| MochaGCN<sub>wo:fusion</sub> | 88.57 | 92.0 | 84.62 |
| MochaGCN<sub>wo:hyperbolic&fusion</sub> | 75.79 | 78.59 | 72.46 |

‘wo’ indicates the removal of a module from the MochaGCN.

## 5 Conclusion

In this paper, we have proposed a novel multimodal hyperbolic graph convolutional neural network for brain network analysis. The proposed method leverages the hyperbolic space to model the intrinsic geometry of the brain network. We conducted experiments on ADNI dataset and demonstrated that the proposed method outperforms baseline methods. The results show that the proposed method achieves a 93.56% classification accuracy with 115 ADNI subjects. This performance not only outperforms baseline methods but also underscores the efficacy of our approach in handling complex neuroimaging data. We also conducted an ablation study to investigate the effect of hyperbolic curvature, embedding dimension, and fusion methods on the performance of the model. The proposed model offers a new perspective in brain network analysis, with significant implications for the diagnosis and treatment of brain disorders. By enhancing the accuracy and efficacy of neuroimaging analyses, our approach holds the potential to greatly improve clinical outcomes for patients with Alzheimer’s disease and other neurodegenerative disorders.

## Supporting information

latex

## Data Availability

All data produced are available online at https://adni.loni.usc.edu/data-samples/adni-data/

## Footnotes

6 https://ida.loni.usc.edu/pages/access/studyData.jsp?project=ADNI

## References

1. Baker, C., Suárez-Méndez, I., Smith, G., Marsh, E.B., Funke, M., Mosher, J.C., Maestú, F., Xu, M., Pantazis, D.: Hyperbolic graph embedding of MEG brain networks to study brain alterations in individuals with subjective cognitive decline. bioRxiv (2023)

2. Cao, X.: Poincaré fréchet mean. Pattern Recognit. 137(109302), 109302 (2023)

3. Chen, T., Kornblith, S., Norouzi, M., Hinton, G.: A simple framework for contrastive learning of visual representations. In: Proceedings of the 37th International Conference on Machine Learning. vol. 119, pp. 1597–1607. PMLR (2020)

4. Febrinanto, F.G., Liu, M., Xia, F.: Balanced graph structure information for brain disease detection. In: Knowledge Management and Acquisition for Intelligent Systems, pp. 134–143. Lecture notes in computer science, Springer Nature Singapore, Singapore (2023)

5. Febrinanto, F.G., Xia, F., Moore, K., Thapa, C., Aggarwal, C.: Graph lifelong learning: A survey. IEEE Computational Intelligence Magazine 18(1), 32–51 (2023)

6. Ganea, O., Bécigneul, G., Hofmann, T.: Hyperbolic neural networks. Advances in neural information processing systems 31 (2018)

7. Lou, A., Katsman, I., Jiang, Q., Belongie, S., Lim, S.N., De Sa, C.: Differentiating through the fréchet mean. In: III, H.D., Singh, A. (eds.) Proceedings of the 37th International Conference on Machine Learning. Proceedings of Machine Learning Research, vol. 119, pp. 6393–6403. PMLR (2020)

8. Meng, X., Liu, J., Fan, X., Bian, C., Wei, Q., Wang, Z., Liu, W., Jiao, Z.: Multi-modal neuroimaging neural network-based feature detection for diagnosis of alzheimer’s disease. Frontiers in Aging Neuroscience 14, 911220 (2022)

9. Nandi, A., Counts, N., Bröker, J., Malik, S., Chen, S., Han, R., Klusty, J., Seligman, B., Tortorice, D., Vigo, D., et al.: Cost of care for alzheimer’s disease and related dementias in the united states: 2016 to 2060. npj Aging 10(1), 13 (2024)

10. Pan, J., Zuo, Q., Wang, B., Chen, C.P., Lei, B., Wang, S.: Decgan: Decoupling generative adversarial network for detecting abnormal neural circuits in alzheimer’s disease. IEEE Transactions on Artificial Intelligence pp. 1–14 (2024)

11. Peng, C., Liu, M., Meng, C., Xue, S., Keogh, K., Xia, F.: Stage-aware brain graph learning for alzheimer’s disease. In: The 2024 IEEE Conference on Artificial Intelligence (CAI) (2024)

12. Peng, C., Liu, M., Meng, C., Yu, S., Xia, F.: Adaptive brain network augmentation based on group-aware graph learning. In: ICLR (2024)

13. Pereira, L.M., Salazar, A., Vergara, L.: A comparative analysis of early and late fusion for the multimodal two-class problem. IEEE Access 11, 84283–84300 (2023)

14. Scheltens, P., De Strooper, B., Kivipelto, M., Holstege, H., Chételat, G., Teunissen, C.E., Cummings, J., van der Flier, W.M.: Alzheimer’s disease. The Lancet 397(10284), 1577–1590 (2021)

15. Selkoe, D.J.: The advent of alzheimer treatments will change the trajectory of human aging. Nature Aging 4(4), 453–463 (2024)

16. Sharpee, T.O.: An argument for hyperbolic geometry in neural circuits. Curr. Opin. Neurobiol. 58, 101–104 (2019)

17. Sun, K., Peng, C., Yu, S., Han, Z., Xia, F.: From electroencephalogram data to brain networks: Graph-learning-based brain disease diagnosis. IEEE Intell. Syst. 39(2), 21–29 (Mar 2024)

18. Whi, W., Ha, S., Kang, H., Lee, D.S.: Hyperbolic disc embedding of functional human brain connectomes using resting-state fMRI. Netw. Neurosci. 6(3), 745– 764 (2022)

19. Xia, F., Sun, K., Yu, S., Aziz, A., Wan, L., Pan, S., Liu, H.: Graph learning: A survey. IEEE Transactions on Artificial Intelligence 2(2), 109–127 (2021)

20. Xu, C., Jia, W., Cui, T., Wang, R., Zhang, Y.f., He, X.: Arbitrary-shape scene text detection via visual-relational rectification and contour approximation. IEEE Transactions on Multimedia 25, 4052–4066 (2023). 10.1109/TMM.2022.3171085

21. Yang, Y., Ye, C., Guo, X., Wu, T., Xiang, Y., Ma, T.: Mapping multi-modal brain connectome for brain disorder diagnosis via cross-modal mutual learning. IEEE Transactions on Medical Imaging (2023)

22. Yu, S., Huang, H., Dao, M.N., Xia, F.: Graph augmentation learning. In: Companion Proceedings of the Web Conference 2022. pp. 1063–1072 (2022)

23. Yu, S., Xia, F., Li, S., Hou, M., Sheng, Q.Z.: Spatio-temporal graph learning for epidemic prediction. ACM Transactions on Intelligent Systems and Technology 14(2), 1–25 (2023)

24. Yue, Y., Lin, F., Yamada, K.D., Zhang, Z.: Hyperbolic contrastive learning. arXiv preprint 2302.01409 (2023)

25. Zeng, L., Li, H., Xiao, T., Shen, F., Zhong, Z.: Graph convolutional network with sample and feature weights for alzheimer’s disease diagnosis. Inf. Process. Manag. 59(4), 102952 (2022)

26. Zhang, H., Rich, P.D., Lee, A.K., Sharpee, T.O.: Hippocampal spatial representations exhibit a hyperbolic geometry that expands with experience. Nat. Neurosci. 26(1), 131–139 (2023)

27. Zhang, J., He, X., Qing, L., Chen, X., Liu, Y., Chen, H.: Multi-relation graph convolutional network for alzheimer’s disease diagnosis using structural MRI. Knowl. Based Syst. 270(110546), 110546 (2023)

28. Zhang, L., Na, S., Liu, T., Zhu, D., Huang, J.: Multimodal deep fusion in hyperbolic space for mild cognitive impairment study. In: Lecture Notes in Computer Science, pp. 674–684. Lecture notes in computer science, Springer Nature Switzerland, Cham (2023)

29. Zhang, Y., He, X., Chan, Y.H., Teng, Q., Rajapakse, J.C.: Multi-modal graph neural network for early diagnosis of alzheimer’s disease from sMRI and PET scans. Comput. Biol. Med. 164(107328), 107328 (2023)

30. Zheng, M., Allard, A., Hagmann, P., Alemán-Gómez, Y., Serrano, M.Á.: Geometric renormalization unravels self-similarity of the multiscale human connectome. Proc. Natl. Acad. Sci. U. S. A. 117(33), 20244–20253 (2020)

31. Zuo, Q., Wu, H., Chen, C.P., Lei, B., Wang, S.: Prior-guided adversarial learning with hypergraph for predicting abnormal connections in alzheimer’s disease. IEEE Transactions on Cybernetics (2024)

