## Supplementary material for "Multimodal Hyperbolic Graph Learning for Alzheimer’s Disease Detection": latex: llncsdoc.pdf

### Instructions for Using Springer's `llncs` Class for Computer Science Proceedings Papers

##### `llncs`, Version 2.24, Jan 29, 2024

#### 1 Installation

Copy `llncs.cls` to a directory that is searched by LaTeX, e.g. either your `texmf` tree or the local work directory with your main LaTeX file.

#### 2 Working with the `llncs` Document Class

##### 2.1 General Information

The `llncs` class is an extension of the standard LaTeX `article` class. Therefore you may use all `article` commands in your manuscript.

If you are already familiar with LaTeX, the `llncs` class should not give you any major difficulties. It basically adjusts the layout to the required standard, defining styles and spacing of headings and captions and setting the printing area to 122 mm horizontally by 193 mm vertically. To keep the layout consistent, we kindly ask you to refrain from using any LaTeX or TeX command that modifies these settings (i.e. `\textheight`, `\vspace`, `baselinestretch`, etc.). Such manual layout adjustments should be limited to very exceptional cases.

In addition to defining the general layout, the `llncs` document class provides some special commands for typesetting the contribution header, i.e. title, authors, affiliations, abstract, and additional metadata. These special commands are described in Sect. 3.

For a more detailed description of how to prepare your text, illustrations, and references, see the *Springer Guidelines for Authors of Proceedings*.

##### 2.2 How to Use the `llncs` Document Class

The `llncs` class is invoked by replacing `article` by `llncs` in the first line of your LaTeX document:

```
\documentclass{llncs}

\begin{document}
  <Your contribution>
\end{document}
```

If your file is already coded with LaTeX, you can easily adapt it to the `llncs` document class by replacing

```
\documentclass{article}
```

with

```
\documentclass{llncs}
```

#### 3 How to Code the Header of Your Paper

##### 3.1 Title

\title Please code the title of your contribution as follows:

```
\title{<Your contribution title>}
```

All words in titles should be capitalized except for conjunctions, prepositions (e.g. on, of, by, and, or, but, from, with, without, under), and definite/indefinite articles (the, a, an), unless they appear at the beginning. Formula letters are typeset as in the text. Long titles that run over multiple lines can be wrapped explicitly with \\. Titles have no end punctuation.

Acknowledgements should generally be placed in an unnumbered subsection at the end of the paper. If you still need to refer to a support or funding program

\thanks in a note to the title, you can use the\thanks macro inside the title:

```
\title{<Your contribution title>\thanks{<granted by x>}}
```

Please do not use \thanks inside \author or \institute as footnotes for these elements are not supported in the online version and will therefore be dropped.

\fnmsep If you need two or more footnotes please separate them with \fnmsep (i.e. *f*oot*n*ote *m*ark *sep*arator).

\titlerunning If a long title does not fit in the single line of the running head, a warning is generated. You can specify an abbreviated title for the running head with the command

```
\titlerunning{<Your abbreviated contribution title>}
```

\subtitle An optional subtitle may also be added:

```
\subtitle{<subtitle of your contribution>}
```

##### 3.2 Author(s)

\author The name(s) of the author(s) are specified by:

```
\author{<author(s) name(s)>}
```

\and If there is more than one author, please separate them by \and. This makes sure that correct punctuation is inserted according to the number of authors.

\inst Numbers referring to different addresses or affiliations should be attached to each author with the \inst{<number>} command. If an author is affiliated with multiple institutions the numbers should be separated by a comma, for example \inst{2,3}.

\orcidID ORCID identifiers can be included with

```
\orcidID{<ORCID identifier>}
```

The ORCID (Open Researcher and Contributor ID) registry provides authors with unique digital identifiers that distinguish them from other researchers and help them link their research activities to these identifiers. Authors who are not yet registered with ORCID are encouraged to apply for an individual ORCID id at https://www.orcid.org and to include it in their papers. In the final publication, the ORCID id will be replaced by an ORCID icon, which will link from the eBook to the actual ID in the ORCID database. The ORCID icon will also replace the number in the printed book.

If you have done this correctly, the author line now reads, for example:

```
\author{First Author\inst{1}\orcidID{0000-1111-2222-3333} \and
Second Author\inst{2,3}\orcidID{1111-2222-3333-4444}}
```

The given name(s) should always be followed by the family name(s). Authors who have more than one family name should indicate which part of their name represents the family name(s), for example by non-breaking spaces `Jos\'{e} Martinez~Perez` or curly braces `Jos\'{e} {Martinez Perez}`.

`\authorrunning` As given name(s) are to be shortened to initials in the running heads, specifying an abbreviated author list with the optional command:

```
\authorrunning{<abbreviated author list>}
```

might add some clarity about the correct representation of author names, in the running-heads as well as in the author index.

##### 3.3 Affiliations

`\institute` Addresses of institutes, companies, etc. should be given in `\institute`.

`\and` Multiple affiliations are separated by `\and`, which automatically assures correct numbering:

```
\institute{<name of an institute>
\and <name of the next institute>
\and <name of the next institute>}
```

`\email` Inside `\institute` you can use

```
\email{<email address>}
```

`\url` and

```
\url{<url>}
```

to provide author email addresses and Web pages. If you need to typeset the tilde character – e.g. for your Web page in your unix system's home directory – the `\homedir` command will do this. If multiple authors have the same affiliation, please check that the order of email addresses matches the sequence of (affiliated) author names.

Please note that, if email addresses are given in your paper, they will also be included in the metadata of the online version.

##### 3.4 Format the Header

`\maketitle` The command `\maketitle` formats the header of your paper. If you leave it out the work done so far will produce *no* text.

##### 3.5 Abstract and Keywords

`abstract` (*env.*) The abstract is coded as follows:

```
\begin{abstract}
<Text of the summary of your paper>
\end{abstract}
```

`\keywords` `\and` Keywords should be specified inside the `abstract` environment. Please capitalize the first letter of each keyword and again separate them with `\and`:

```
\keywords{First keyword \and Second keyword \and Third keyword}
```

The keyword separator will then be properly rendered as a middle dot.

#### 4 How to Code the Body of Your Paper

##### 4.1 General Rules

From a technical point of view, the `llncs` document class does not require any specific LaTeX coding in the body of your paper. You can simply use the commands provided by the 'article' document class. For more information about what will be done with your manuscript before publication, please refer to the *Springer Guidelines for Authors of Proceedings*.

##### 4.2 Special Math Characters

The `llncs` document class supports some additional special characters:

| | | | | | |
|---|---|---|---|---|---|
| `\grole` | yields | $\gtrless$ | `\getsto` | yields | $\leftrightarrows$ |
| `\lid` | yields | $\leqq$ | `\gid` | yields | $\geqq$ |

If you need blackboard bold characters, i.e. for sets of numbers, please load the related $\mathcal{A}\mathcal{M}\mathcal{S}$-TEXfonts. If for some reason this is not possible you can also use the following commands from the `llncs` class:

| | | | | | |
|---|---|---|---|---|---|
| `\bbbc` | yields | $\mathbb{C}$ | `\bbbf` | yields | $\mathbb{F}$ |
| `\bbbh` | yields | $\mathbb{H}$ | `\bbbk` | yields | $\mathbb{K}$ |
| `\bbbm` | yields | $\mathbb{M}$ | `\bbbn` | yields | $\mathbb{N}$ |
| `\bbbp` | yields | $\mathbb{P}$ | `\bbbq` | yields | $\mathbb{Q}$ |
| `\bbbr` | yields | $\mathbb{R}$ | `\bbbs` | yields | $\mathbb{S}$ |
| `\bbbt` | yields | $\mathbb{T}$ | `\bbbz` | yields | $\mathbb{Z}$ |
| `\bbbone` | yields | $\mathbb{1}$ | | | |

Please note that all these characters are only available in math mode.

### 5 Theorems, Definitions, and Proofs

#### 5.1 Predefined Theorem-Like Environments

`corollary` (*env.*)
`definition` (*env.*)
`lemma` (*env.*)
`proposition` (*env.*)
`theorem` (*env.*)

Several theorem-like environments are predefined in the `llncs` document class. The following environments have a bold run-in heading, while the following text is in italics:

```
\begin{corollary}   <text> \end{corollary}
\begin{definition}  <text> \end{definition}
\begin{lemma}       <text> \end{lemma}
\begin{proposition} <text> \end{proposition}
\begin{theorem}     <text> \end{theorem}
```

`case` (*env.*)
`conjecture` (*env.*)
`example` (*env.*)
`exercise` (*env.*)
`note` (*env.*)
`problem` (*env.*)
`property` (*env.*)
`question` (*env.*)
`remark` (*env.*)
`solution` (*env.*)

Other theorem-like environments render the text in roman, while the run-in heading is bold as well:

```
\begin{case}        <text> \end{case}
\begin{conjecture}  <text> \end{conjecture}
\begin{example}     <text> \end{example}
\begin{exercise}    <text> \end{exercise}
\begin{note}        <text> \end{note}
\begin{problem}     <text> \end{problem}
\begin{property}    <text> \end{property}
\begin{question}    <text> \end{question}
\begin{remark}      <text> \end{remark}
\begin{solution}    <text> \end{solution}
```

`claim` (*env.*)
`proof` (*env.*)

Finally, there are also two unnumbered environments that have the run-in heading in italics and the text in upright roman.

```
\begin{claim}       <text> \end{claim}
\begin{proof}       <text> \end{proof}
```

`\qed`

Proofs may contain an eye catching square, which can be inserted with `\qed`) before the environment ends.

#### 5.2 User-Defined Theorem-Like Environments

`\spnewtheorem`

We have enhanced the standard `\newtheorem` command and slightly changed its syntax to get two new commands `\spnewtheorem` and `\spnewtheorem*` that now can be used to define additional environments. They require two additional arguments, namely the font style of the label and the font style of the text of the new environment:

```
\spnewtheorem{<env_nam>}[<num_like>]{<caption>}{<cap_font>}{<body_font>}
```

For example,

```
\spnewtheorem{maintheorem}[theorem]{Main Theorem}{\bfseries}{\itshape}
```

will create a *main theorem* environment that is numbered together with the predefined *theorem*. The sharing of the default counter ([theorem]) is desired. If you omit the optional second argument of \spnewtheorem, a separate counter for your new environment is used throughout your document.

In combination with the (obsolete) class option envcountsect (see. Sect. 8), the \spnewtheorem command also supports the syntax:

```
\spnewtheorem{<env_nam>}{<caption>}[<within>]{<cap_font>}{<body_font>}
```

With the parameter <within>, you can control the sectioning element that resets the theorem counters. If you specify, for example, subsection, the newly defined environment is numbered subsectionwise.

\spnewtheorem* If you wish to add an unnumbered environment, please use the syntax

```
\spnewtheorem*{<env_nam>}{<caption>}{<cap_font>}{<body_font>}
```

#### 6 Credits and Acknowledgments

credits (*env.*) Credits and acknowledgments should be placed at the end of the paper, just before the references. Please use the credits environment to make sure that both text and run-in headings are printed in small font size.

\ackname
\discintname
There are two possible \subsubsection headings to provide such statements: "Acknowledgments" and "Disclosure of Interests". The \ackname and \discintname macros are used to generate the correct run-in titles.

General acknowledgments can be provided in the (optional) paragraph "Acknowledgments", which is followed by the (mandatory) paragraph "Disclosure of Interests", where any competing interests of the authors are to be declared.

#### 7 References

There are three options for citing references:

- arabic numbers, i.e. [1], [3–5], [4–6,9],
- labels, i.e. [CE1], [AB1,XY2],
- author/year system, (Smith et al. 2000), (Miller 1999a, 12; Brown 2018).

We prefer citations with arabic numbers, i.e. the usage of \bibitem without an
citeauthoryear optional parameter. If you want to use the author/year system, you can use the class option citeauthoryear, i.e.

```
\documentclass[citeauthoryear]{llncs}
```

Please note that this option does not automatically change your citations to the author/year style. It basically redefines the \bibitem command to take the publication year as an optional parameter that is displayed instead of an arabic number. Author name(s) and, if necessary, parentheses are to be typed manually. If your reference reads

```
\bibitem[2016]{vdaalst:2016}
van der Aalst, W.: Process Mining, 2nd ed. Springer, Heidelberg (2016)
```

and is cited as follows:

```
... is shown by van der Aalst (\cite{vdaalst:2016})
```

the resulting text will be:

> "...is shown by van der Aalst (2016)."

splncs04.bst We encourage you to use BibTeX for typesetting your references. For formatting the bibliography according to Springer's standard (for mathematics, physical sciences, and computer science), please use the bibliography style file splncs04.bst that comes with the llncs document class. You simply need to add \bibliographystyle{splncs04} to your document. DOIs should be provided in the doi field of your .bib database. BibTeX will then automatically add them to your references.

\doi If you do not use BibTeX, you can include a DOI with the \doi command:

```
\doi{<DOI>}
```

The DOI will be expanded to the URL https://doi.org/<DOI> in accordance with the CrossRef guidelines.

#### 8 Obsolete Class Options

The llncs document class contains several class options that have become obsolete over the years. We only mention them for completeness:

- orivec – The llncs document class changes the formatting of vectors coded with \vec to boldface italics. If you absolutely need the original LaTeX design for vectors, i.e. an arrow above the related variable, you can restore it with the orivec option.
- envcountsame – All theorem-like environments share one counter, i.e. Theorem 1, Lemma 2, Corollary 3, etc.
- envcountreset – All theorem-like environments are numbered per section, i.e. the related counters are reset to 1 in every section.
- envcountsect – All theorem-like environments are numbered per section, and the section number added to the individual counter, i.e. Theorem 1.2, Lemma 2.2, etc.
- openbib – This option produces the "open" bibliography style, in which each block starts on a new line, and succeeding lines in a block are indented by \bibindent.
- oribibl – This option restores the original LaTeX definitions for the bibliography and the \cite mechanism that some BibTeX applications rely on.
