## Supplementary material for "Multimodal Hyperbolic Graph Learning for Alzheimer’s Disease Detection": latex: MochaGCN.tex

\documentclass[runningheads]{llncs}

\usepackage[T1]{fontenc}
\usepackage{graphicx}
\usepackage{subcaption}
\usepackage[export]{adjustbox}
\usepackage{orcidlink}
\usepackage[misc]{ifsym}
\usepackage{cite}
\usepackage{amsmath}
\usepackage{amssymb}
\usepackage{colortbl}
\usepackage{ctable}
\usepackage{booktabs,caption}
\usepackage[flushleft]{threeparttable}
\usepackage{array}
\usepackage{multicol}
\usepackage{tikz}
\usepackage{pgfplots}
\usepackage{filecontents}
\usetikzlibrary{patterns}
\usepackage{algorithm,algorithmic}
\usepackage{fixltx2e}

\hypersetup{
    colorlinks=false,
    pdfborder={0 0 0},
}
\captionsetup[table]{skip=6pt}

\definecolor{color1}{rgb}{0.98, 0.81, 0.69}
\definecolor{color2}{rgb}{0.55, 0.71, 0.0}
\definecolor{color3}{rgb}{1.0, 0.6, 0.4}
\definecolor{color4}{rgb}{0.29, 0.59, 0.82}
\definecolor{color5}{HTML}{b4b4b4}
\definecolor{color6}{HTML}{5f97d2}
\definecolor{color7}{HTML}{c76d72}
\definecolor{bblue}{HTML}{4F81BD}
\definecolor{rred}{HTML}{C0504D}
\definecolor{ggreen}{HTML}{9BBB59}
\definecolor{ppurple}{HTML}{9F4C7C}
\definecolor{dred}{HTML}{D9474C}
\definecolor{dorange}{HTML}{FFA964}
\definecolor{dyellow}{HTML}{FFDC9D}
\definecolor{dgreen}{HTML}{64C4AB}
\definecolor{dblue}{HTML}{68C6E0}
\definecolor{ddblue}{HTML}{41609C}
\definecolor{ddgreen}{HTML}{93ce97}
\definecolor{ddpurple}{HTML}{ef8c8e}

\DeclareMathOperator\arctanh{arctanh}
\DeclareMathOperator\arccosh{arccosh}

\def\curvature{-1.0}
\def\dimension{128}
\def\acc{93.56}
\def\sen{95.34}
\def\spe{91.54}
\def\algorithmName{MochaGCN}


\begin{document}


\title{Multimodal Hyperbolic Graph Learning for Alzheimer's Disease Detection}
\titlerunning{Multimodal Hyperbolic Graph Learning for Alzheimer's Disease Detection}


\author{Chengyao Xie\inst{1}\orcidlink{0009-0004-9111-2127} \and
Wenhao Zhou\inst{2}\orcidlink{0009-0000-0413-4282} \and
Ciyuan Peng\inst{1}\orcidlink{0000-0002-3580-5402} \and
Azadeh Noori Hoshyar \inst{1}\orcidlink{0000-0001-7729-8907} \and
Chengpei Xu \inst{3}\orcidlink{0000-0001-8860-7701} \and
Usman Naseem \inst{4}\orcidlink{0000-0003-0191-7171} \and
Feng Xia\inst{5}\textsuperscript{(\Letter)}\orcidlink{0000-0002-8324-1859} }
\authorrunning{C. Xie et al.}

\institute{
    Institute of Innovation, Science and Sustainability, Federation University Australia, Ballarat, VIC 3353, Australia \and
    School of Internet, Anhui University, Hefei, 230039, China \and
    MIoT \& IPIN Lab, School of Minerals and Energy Resources Engineering, University of New South Wales, Sydney, NSW 2052, Australia \and
    School of Computing, Macquarie University, Sydney, NSW 2109, Australia \and
    School of Computing Technologies, RMIT University, Melbourne, VIC 3000, Australia \\
    \{chengyao.xie, ciyuan.peng, a.noorihoshyar\}@federation.edu.au,
}
%


\maketitle
\begin{abstract}
Multimodal graph learning techniques have demonstrated significant potential in modeling brain networks for Alzheimer's disease (AD) detection. However, most existing methods rely on Euclidean space representations and overlook the scale-free and small-world properties of brain networks, which are characterized by power-law distributions and dense local clustering of nodes. This oversight results in distortions when representing these complex structures. To address this issue, we propose a novel multimodal Poincaré Fréchet mean graph convolutional network (MochaGCN) for AD detection. MochaGCN leverages the exponential growth characteristics of hyperbolic space to capture the scale-free and small-world properties of multimodal brain networks. Specifically, we combine hyperbolic graph convolution and Poincaré Fréchet mean to extract features from multimodal brain networks, enhancing their representations in hyperbolic space. Our approach constructs multimodal brain networks by integrating information from diffusion tensor imaging (DTI) and functional magnetic resonance imaging (fMRI) data. Experiments on the Alzheimer's Disease Neuroimaging Initiative (ADNI) dataset demonstrate that the proposed method outperforms state-of-the-art techniques.

\keywords{Hyperbolic space \and Graph learning \and Graph convolutional networks \and Multimodal fusion \and Alzheimer’s disease.}
\end{abstract}


% --------------------- Introduction ----------------------------
\section{Introduction}
Alzheimer's disease (AD) is an irreversible neurodegenerative disorder that impacts cognition, function, and behavior, progressively leading to a loss of physical functions and, ultimately, death \cite{scheltens2021alzheimer}. Approximately 60 million people worldwide were affected by Alzheimer's disease \cite{selkoe2024advent}, and the global economic burden is projected to reach 9.12 trillion US dollars by 2050 \cite{nandi2024cost}. These staggering figures underscore the urgent need for more effective diagnostic and monitoring tools. The disease is linked with the accumulation of amyloid beta and tau proteins in specific cortex areas, causing the loss of neurons and synapses. Neuroimaging that reflects the structural, functional connectivity properties of the human brain can be used to analyze disease development.


Recently, graph learning techniques have demonstrated significant capabilities in brain network modeling, achieving promising results in brain disorder detection \cite{Xia2021, Sun2024-fd, yu2023spatio, yu2022graph, febrinanto2023graph }. With the increasing availability of multimodal neuroimaging data, such as functional magnetic resonance imaging (fMRI), diffusion tensor imaging (DTI), computed tomography (CT), and positron emission tomography (PET), multimodal brain graph learning has become a trending research area. Some works enhance the detection performance by integrating graph neural network (GNN) and its variants with novel multimodal feature learning \cite{Peng2024-ws, Zeng2022-kn}. In addition, some studies highlight that the graph structure information is crucial for advancing the brain disorder detection performance \cite{peng2024adaptive, Febrinanto2023-wt, Zhang2023-el}. For example, Zhang et al. \cite{Zhang2023-yw} proposed a multimodal graph neural network to fuse the learned node representations for AD prediction. However, these methods rely on Euclidean space representations, which are not suitable for capturing the scale-free and small-world properties of brain networks \cite{Zheng2020-tw, Zhang2023-vq}, as shown in Fig. \ref{fig:image0}. As brain regions are organized hierarchically, the regions and their connections grow exponentially. This exponential growth surpasses the polynomial expansion capacity of Euclidean space, leading to \textbf{representation distortion}.

Notably, hyperbolic geometry can describe data structures that grow exponentially more effectively than Euclidean space \cite{Zheng2020-tw, Sharpee2019-nb}.
Recent studies use hyperbolic graph learning to achieve superior representations and improve model availability \cite{Zhang2023-lw, Baker2023-zg, xu2022, Whi2022-za}. For example, Zhang et al. \cite{Zhang2023-lw}, projected the functional and structural brain network into hyperbolic space and utilized a hyperbolic graph kernel for the diagnosis of mild cognitive impairment (MCI). These works demonstrate that hyperbolic geometry can better capture graph structures and improve model performance.


Therefore, to reduce representation distortion of multimodal brain networks and enhance information fusion, we propose a novel multimodal hyperbolic graph learning method (\algorithmName) for AD detection. Specifically, we introduce a hyperbolic graph convolutional network with Poincaré Fréchet mean aggregation to capture multimodal brain network features in hyperbolic space. We fuse information from DTI and fMRI modalities and employ contrastive learning to enhance the fused features by minimizing the contrastive loss. Finally, we calculate the hyperbolic distance of the learned representation features for AD detection. The proposed model offers a more accurate representation of multimodal brain networks, holding significant implications for the diagnosis and treatment of brain disorders.


\vspace{0.3em}
Our contributions can be summarized as follows:
\vspace{-0.3em}
\begin{itemize}
    \item[$-$] We propose a novel multimodal hyperbolic Poincaré Fréchet mean graph convolutional neural network (\algorithmName) for AD detection, significantly reducing the representation distortion of brain networks and enhancing information fusion between DTI and fMRI modalities.
    \vspace{0.3em}
    \item[$-$] We combine a graph convolutional network with the Poincaré Fréchet mean aggregation to preserve scale-free and small-world properties of multimodal brain network representations in hyperbolic space.
    \vspace{0.3em}
    \item[$-$] The experimental results on the Alzheimer’s Disease Neuroimaging Initiative (ADNI) dataset demonstrate the effectiveness of the proposed method, which outperforms the state-of-the-art methods.
\end{itemize}

\begin{figure}[ht]
    \hspace{0.05\textwidth}
    \begin{minipage}[b]{0.28\linewidth}
        \centering
        \includegraphics[width=\linewidth,valign=c]{figure/scale_free.png}
        (a) Brain network node degree distribution
    \end{minipage}
    \hspace{0.05\textwidth}
    \begin{minipage}[b]{0.25\linewidth}
        \centering
        \includegraphics[width=\linewidth,valign=c]{figure/fig08.png}
        (b) DTI network connections
    \end{minipage}
    \hspace{0.05\textwidth}
    \begin{minipage}[b]{0.25\linewidth}
        \centering
        \includegraphics[width=\linewidth,valign=c]{figure/fig07.png}
        (c) fMRI network connections
    \end{minipage}

    \caption{Scale-free and small-world properties of brain networks. (a) The node degree distribution of the fMRI brain networks from 115 subjects in the ADNI dataset follows a power-law distribution, indicates the scale-free characteristics. The visualizations of (b) the DTI network and (c) the fMRI network projected onto the Poincaré disk for one subject demonstrate small-world characteristics.}
    \label{fig:image0}
\end{figure}


% ------------------- Introduction end --------------------------


% --------------------- Preliminaries ----------------------------
\section{Preliminaries}


\subsection{Problem Definition}
The AD detection task first obtains the connectivity from preprocessed DTI and fMRI neuroimages to construct the brain networks. The networks are represented as graph \( \mathcal{G}_{\mathrm{DTI}} = (V_{\mathrm{DTI}}, E_{\mathrm{DTI}}) \) and \( \mathcal{G}_{\mathrm{fMRI}} = (V_{\mathrm{fMRI}}, E_{\mathrm{fMRI}}) \), where \( V \) is the set of vertices representing brain regions and \( E \) is the set of edges connecting the regions. The connectivity graph in Euclidean space is represented by an adjacency matrix \({\mathbf{A}}_{\mathrm{DTI}}^E, {\mathbf{A}}_{\mathrm{fMRI}}^E\in \mathbb{R}^{n \times n}\), where \(n\) is the number of regions of interest (ROIs). Each element \( e_{i,j} \) of \(\mathbf{A}\) represents the connectivity strength between regions \( i \) and \( j \). Each modality's graphs form the data \({\mathbf{M}_{\mathrm{DTI}}}\) and \({\mathbf{M}_{\mathrm{fMRI}}}\), where \(\mathbf{M} = \{\mathbf{A}_1,\mathbf{A}_2,...,\mathbf{A}_{n_p} \}\) and \(n_p\) is the number of participants. The goal of this research is to learn a classification function \( f: (\mathbf{A}_{\mathrm{DTI}}^{E}, \mathbf{A}_{\mathrm{fMRI}}^{E}) \rightarrow p  \), where \( p\) is the probability of subject class $\mathbf{\gamma} $ , i.e., \( \gamma = 1 \) for AD patients and \( \gamma = 0 \) for normal controls. The function \( f(\cdot) \) is learned from a training set of subjects with known labels. The learned function is then used to predict the labels of new subjects in a test set and its performance is evaluated using standard classification metrics.


\subsection{Hyperbolic Geometry}
Hyperbolic geometry, characterized by negative curvature, differs from the flat Euclidean space. 
Hyperbolic space grows exponentially. Specifically, considering a 2-dimensional hyperbolic disk with radius $r$ and constant curvature -1, the perimeter and area of the disk can be calculated by $2\pi\sinh r$ and $2\pi(\cosh r - 1)$, and both of them grow as $e^r$ with $r$, compared with the linearly and quadratically growth in Euclidean space. So it is ideal for representing data with exponential expansion and hierarchical structures. 
There are many models to embed hyperbolic space in Euclidean space. And in this paper, the Poincaré model is introduced as the necessary background of this study. Hyperbolic manifold \(\mathbb{H}\) with curvature \(K < 0\) can be mapped to  Euclidean space by the Poincaré ball model, which is defined as:
\begin{equation}
    \mathbb{H}_K^d:=\{\phi\in \mathbb{R}^d: \|\phi\|^2 < -\frac{1}{K}\},
\end{equation}
where \(\|\cdot\|\) denotes the Euclidean norm. When \(\lim K = 0\), the hyperbolic space is isometric to the Euclidean space. For two points \(\phi_1, \phi_2\) in the hyperbolic space, the M\"obius addition is defined as:
\begin{equation}
    \phi_1\oplus_K \phi_2\!\! =\!\! \frac{(1 \!-\! 2K\langle \phi_1,\!\phi_2\rangle\!-\!K\|\phi_2\|^2)\phi_1 \!+\! (1\!+\!K\|\phi_1\|^2)\phi_2}{1 - 2K\langle \phi_1,\!\phi_2 \rangle + K^2 \|\phi_1\|^2 \|\phi_2\|^2}.
\end{equation}
And the distance between them is defined as
\begin{equation}\label{hyperbolic_distance}
    d^{K}(\phi_1,\phi_2)=\frac{2}{\sqrt{|K|}}\mathrm{arctanh}(\sqrt{|K|}\,||-{\phi_1}\oplus_{K}{\phi_2}||).
\end{equation}

For a point \(\phi\) in the hyperbolic space, the tangent space \(\mathcal{T}\mathbb{H}_K^d\) at \(\phi\) is Euclidean space. The coordinates can be mapped to each other by the exponential map and logarithmic map. The exponential map \({exp}_{K}: \mathcal{T}\mathbb{H}_K^d \rightarrow \mathbb{H}_K^d\) maps a vector \({\phi^E}\) in the tangent space to a point in the hyperbolic space, and the logarithmic map \({log}_{\phi}^{K}: \mathbb{H}_K^d \rightarrow \mathcal{T}\mathbb{H}_K^d\) maps a point \({\phi^H}\) in the hyperbolic space to a vector in the tangent space. For a given point $\phi$, the exponential map of point \({\phi^E}\) and logarithmic map of point \({\phi^H}\) are defined as:
\begin{equation}\label{eq_exp_map}
    {exp}_{K}({\phi^E}):={\phi}\oplus_K\Big(\tanh(\sqrt{|K|}\frac{\lambda_{K} \|{\phi^E}\|}{2})\frac{{\phi^E}}{\sqrt{|K|}\,\|{\phi^E}\|}\Big),
\end{equation}
\begin{equation}\label{eq_log_map}
    {log}_{K}({{\phi^H}}):=\frac{2}{\sqrt{|K|\lambda_{K}}}\arctanh(\sqrt{|K|}\,\|-{\phi}\oplus_{K}{{\phi^H}}\|)\frac{-{\phi}\oplus_{K}{{\phi^H}}}{||-{\phi}\oplus_{K}{{\phi^H}}||},
\end{equation}
where \(\lambda_{K} = 2(1+K\|\phi\|^2)^{-1}\) is the conformal factor.


According to \cite{Ganea2018-ct}, we formulate matrix-vector multiplication, bias translation, and activation function of hyperbolic neural networks are defined as:
\begin{equation}
    \mathbf{T} \otimes_{K} \mathbf{\omega} = \exp_{K}(\mathbf{T} \log_{K}(\mathbf{\omega})),
\end{equation}
\begin{equation}
    \mathbf{\omega}\oplus_{K} \mathbf{b}=\exp_{K}(P\mathbf{T}_{K}(\mathbf{b})),
\end{equation}
\begin{equation}\label{eq_activation}
    \sigma_{K_{1}, K_{2}}(\mathbf{\omega}) = \exp_{K_{1}}(\sigma(\log_{K_{2}}(\mathbf{\omega}))),
\end{equation}
respectively, where \(\mathbf{T}\) is a matrix, \(\mathbf{\omega}\) is a vector, \(\mathbf{b}\) is a bias, and \(\sigma\) is an activation function.
% ------------------- Preliminaries end --------------------------


% --------------------- Methodology ----------------------------
\section{Methodology}

The AD detection task is a binary classification problem and involves four main steps. First, functional, structural, and fused brain networks are constructed for each subject based on DTI and fMRI imaging data. Next, hyperbolic graph convolution is applied to these brain networks to extract representation feature vectors. Then, contrastive learning minimizes the contrastive loss between the fused vector and the functional and structural vectors. Finally, the fused feature vectors are projected into hyperbolic space, and the distance from the cluster center is calculated to generate the diagnosis results. The overall architecture of the proposed method is shown in Fig. \ref{fig:architecture}.

\begin{figure}[h]
    \centering
    \includegraphics[width=0.98\linewidth]{figure/framework.png}
    \caption{Overall framework of \algorithmName. (a) Calculation of the fMRI functional connectivity matrix and DTI structural connectivity matrix based on the AAL brain atlas, followed by graph construction. (b) Projection of the brain graph to hyperbolic space and fusion of the DTI fMRI information into a fusion modality, with hyperbolic Poincaré Frechet mean graph convolution (Graph Conv.) applied to each modality. (c) Feature learning for DTI and fMRI modalities via contrastive learning, where the fusion feature is learned by contrasting the fusion feature with the other two features. (d) Disease classification by calculating the fusion feature's hyperbolic distance.}
    \label{fig:architecture}
\end{figure}

\subsection{Brain Graph Construction}
We begin by preprocessing the neuroimaging data to construct structural graph $\mathbf{A}_{\mathrm{DTI}}^{E}$ derived from DTI data, and functional graph $\mathbf{A}_{\mathrm{fMRI}}^{E}$ based on fMRI data. Both graphs are initially represented in Euclidean space. Subsequently, we employ the mapping techniques outlined in Section \ref{mapping} to project these graphs into hyperbolic space, and get hyperbolic counterparts $\mathbf{A}_{\mathrm{DTI}}^H$, $\mathbf{A}_{\mathrm{fMRI}}^H$.

We introduce a multimodal fusion graph to leverage both early and late fusion strategies \cite{Pereira2023-dc}. In the early fusion stage, we integrate the information from both modalities in hyperbolic space. This fusion is formulated as follows:
\begin{equation}\label{eq:fusion}
    \mathbf{A}_{\mathrm{fusion}}^H=\mathbf{I}\oplus_{K}\left(\alpha\otimes_{K}\mathbf{A}_{\mathrm{DTI}}^H\right)\oplus_{K}\left(\beta\otimes_{K}\mathbf{A}_{\mathrm{fMRI}}^H\right),
\end{equation}
where $\mathbf{I}$ is the identity matrix. $\alpha,\beta$ are the fusion parameters, and $\alpha+\beta = 1$. 

\subsection{Hyperbolic Graph Convolution}
To capture the brain graph features for AD detection, a hyperbolic graph convolutional network will be used to learn the representation of the brain network. The input graphs are represented by a binary adjacency matrix and the outputs are hyperbolic embeddings for graph classification and link prediction tasks. The proposed model architecture is illustrated in Fig. \ref{fig:architecture}. The model consists of the following parts:
\begin{itemize}
    \item[$-$]\textbf{Mapping} \label{mapping} We first transport the input data between Euclidean space and hyperbolic space via logarithmic map Eq. (\ref{eq_log_map}) and exponential map Eq. (\ref{eq_exp_map}). For DTI and fMRI brain networks, we first map the input data to hyperbolic space using the exponential map $ \mathbf{A}_{\mathrm{DTI}}^H=exp_{K}(\mathbf{A}_{\mathrm{DTI}}^{E})$ and $ \mathbf{A}_{\mathrm{fMRI}}^H=exp_{K}(\mathbf{A}_{\mathrm{fMRI}}^{E})$, where $\mathbf{A}^{H}$ represents the brain graph in hyperbolic space.

\vspace{0.5em}
\item[$-$]\textbf{Node Aggregation} Aggregation function captures features of neighborhood in the graph, and is a core component of convolutional networks. Lou et al. \cite{pmlr-v119-lou20a} extended Fréchet mean for arbitrary Riemannian manifolds, and Cao \cite{Cao2023-hm} presented a ($1-\epsilon$)-approximation to fix imprecise convergences problem. In this work, we combine the Poincaré Fréchet mean and hyperbolic graph convolution to aggregate the information from neighboring nodes. Each modality adjacent matrix in hyperbolic space, $\mathbf{A}_{\mathrm{DTI}}^{H}$, $\mathbf{A}_{\mathrm{fMRI}}^{H}$, $\mathbf{A}_{\mathrm{fusion}}^{H}$ is then fed into the hyperbolic graph convolution module to learn the corresponding representations. This process is formulated as:
\begin{equation}\label{eq:weight}
    \mathbf{h}_{i}^{\ell}=\mathbf{W}^{\ell}\otimes_{K_{\ell-1}}\mathbf{x}_{i}^{\ell-1}\oplus_{K_{\ell-1}}\,\mathbf{b}^{\ell},
\end{equation}
\begin{equation}
    \mathbf{y}_{i}^{\ell}=\mathrm{Aggregation}_{j\in N(i)}^{{\mu}_{\mathrm{PF}}}(\mathbf{h}_{j}^{\ell}),
\end{equation}
where ${\ell}$ is the layer index, where $\mathbf{x}_{i}^{\ell-1}$ are the features of node $i$ of the previous layer
$\ell-1$, and $\mathbf{W}^{\ell}$ and $\mathbf{b}^{\ell}$ the weights and bias parameters of layer ${\ell}$, $N(i)$ denotes the neighbors of node $i$. The Poincaré Fréchet mean ${\mu}_{\mathrm{PF}}$ is defined as:
\begin{equation}
    {\mu}_{\mathrm{PF}}={\arg \min } \sum_{k=1} \mathbf{w}_k \cdot d^K\left(\mathbf{x}_{k}, \mu\right)^2,
\end{equation}
where $d^K$ is the the hyperbolic distance as introduced in Eq. (\ref{hyperbolic_distance}), $w_k$ is the weight of the $k$-th point, and $\mu$ is the Fréchet mean of the points in the manifold.
\vspace{0.5em}
\item[$-$]\textbf{Activation Function} We implement the hyperbolic ReLU function using Eq. (\ref{eq_activation}). The output was defined by: 
\begin{equation}\label{eq:Activation}
    \mathbf{x}^{l}_{i}=\sigma(\mathbf{y}^{l}_{i}).
\end{equation}

\end{itemize}

The output of each modality is $\mathbf{x}^{l}_{\mathrm{DTI}}$, $\mathbf{x}^{l}_{\mathrm{fMRI}}$, and $\mathbf{x}^{l}_{\mathrm{fusion}}$ respectively, where $l$ is the number of layers. 


\subsection{Contrastive Fusion}
According to \cite{Pereira2023-dc}, we will leverage late contrastive fusion techniques, further enhancing the fusion modality features based on previously learned representations $\mathbf{x}^{l}_{\mathrm{DTI}}$, $\mathbf{x}^{l}_{\mathrm{fMRI}}$, and $\mathbf{x}^{l}_{\mathrm{fusion}}$. For the DTI modality, we select subjects with the same class labels as positive samples and those with different class labels as negative samples from the dataset and follow the approach of SimCLR \cite{Chen2020-au} and HCL \cite{Yue2023-gq} to construct a contrastive loss, as follows:
\begin{equation}\label{eq:loss_dti}
    \mathcal{L}_{\mathrm{DTI}} = -\sum_{i\in N}\log\left({\frac{\exp{\big(}-d^K(\mathbf{x}_\mathrm{i,DTI}^{l},\mathbf{x}_\mathrm{pos(i),DTI}^{l})/\tau\big)}{\sum_{a\in M}\exp{\big(}-d^K(\mathbf{x}_\mathrm{i,DTI}^{l},\mathbf{x}_\mathrm{a,DTI}^{l})/\tau{\big)}}}\right),
\end{equation}
where $\tau\in{R^+}$ is a scalar temperature parameter, $M$ denotes a batch samples, $\mathbf{x}_{pos(i)}$ is the positive pairs of node $i$. We can also obtain a contrastive loss $\mathcal{L}_{\mathrm{fMRI}}$ for fMRI modality in a similar way. These two contrastive losses are used to update the DTI and fMRI representations separately without exchanging information with each other.

Since the fused modality integrated the information from both modalities early on, it will further refine this information by comparing to the features extracted from DTI and fMRI modalities in the late stage. The fusion modality contrastive loss can be calculated by:
\begin{equation}\label{eq:loss_fusion}
    \begin{split}
    \mathcal{L}_{\mathrm{fusion}} = -\sum_{i\in M}\log\left({\frac{\exp{\big(}-d^K(\mathbf{x}_\mathrm{i,fusion}^{l},\mathbf{x}_\mathrm{pos(i),DTI}^{l})/\tau\big)}{\sum_{a\in M}\exp{\big(}-d^K(\mathbf{x}_\mathrm{i,fusion}^{l},\mathbf{x}_\mathrm{a,DTI}^{l})/\tau{\big)}}}\right) \\
    -\sum_{i\in M}\log\left({\frac{\exp{\big(}-d^K(\mathbf{x}_\mathrm{i,fusion}^{l},\mathbf{x}_\mathrm{pos(i),fMRI}^{l})/\tau\big)}{\sum_{a\in M}\exp{\big(}-d^K(\mathbf{x}_\mathrm{i,fusion}^{l},\mathbf{x}_\mathrm{a,fMRI}^{l})/\tau{\big)}}}\right).
    \end{split}
\end{equation}

\subsection{Classification}
The AD classification probability is calculated using the hyperbolic distance between the fusion modality's extracted features and the centers $ \mathcal{C} = [ \mathbf{c}_1, \mathbf{c}_2, ..., \mathbf{c}_{q} ] $ of different classes, where $q$ is the number of classes. The probability of the $j$-th class is calculated as follows:
\begin{equation}\label{eq:classification}
    p(\gamma_j) = \textrm{softmax} \left( \mathbf{\psi}_{0},..., \mathbf{\psi}_{q} \right),
\end{equation}
where $\psi_{i} = (\exp({d^{K}(\mathbf{x}_{\mathrm{fusion}},\mathbf{c}_{i})^{2}-2}) + 1)^{-1}$ and $\gamma_j$ is the $j$-th class. 


To elucidate the data processing mechanism of \algorithmName, we employ pseudocode to describe the model's AD classification process, as outlined in Algorithm 1.
\begin{algorithm}[t]
    \renewcommand{\algorithmicrequire}{\textbf{Input:}}
    \renewcommand{\algorithmicensure}{\textbf{Output:}}
    \caption{Learning process of \algorithmName}
    \label{alg:pipeline}
    \begin{algorithmic}[1]
        \REQUIRE A training epoch $g$ of multimodal data ${\mathbf{M}_{\mathrm{fMRI}}},{\mathbf{M}_{\mathrm{DTI}}}$, where ${\mathbf{A}}_i \in {M}$
        \ENSURE AD classification result ${p(\gamma)}$
        \FOR{$i = 1, 2, \cdots, g$}
            \STATE Map the DTI and fMRI graph into hyperbolic space $\mathbf{A}_\mathrm{i,DTI}$, $\mathbf{A}_\mathrm{i,fMRI}$ (Eq. (\ref{eq_exp_map}), Eq. (\ref{eq_log_map})) and construct fusion modality graph $\mathbf{A}_\mathrm{i,fusion}$ (Eq. (\ref{eq:fusion}))
            \STATE Extract each modality's features by hyperbolic graph convolution and get $\mathbf{x}_\mathrm{i,DTI}$, $\mathbf{x}_\mathrm{i,fMRI}$, $\mathbf{x}_\mathrm{i,fusion}$ (Eq. (\ref{eq:weight})-Eq. (\ref{eq:Activation}))
            \STATE Calculate the contrastive loss $\mathcal{L}_{\mathrm{i,DTI}}$, $\mathcal{L}_{\mathrm{i,fMRI}}$ and $\mathcal{L}_{\mathrm{i,fusion}}$ between the positive and negative samples (Eq.(\ref{eq:loss_dti}), Eq. (\ref{eq:loss_fusion})) 
            \STATE Update each modality's neural network parameters by minimizing the contrastive loss
        \ENDFOR
        \STATE Calculate the AD classification result ${p(\gamma)}$ (Eq. (\ref{eq:classification}))
    \end{algorithmic}
\end{algorithm}

\subsection{Complexity Analysis}

The time complexity of the primary component of \algorithmName\ is analyzed below. The complexity for mapping the brain graph to hyperbolic and tangent spaces is ${O}(n^2)$, where $n$ represents the number of ROIs. The complexity for forward training and backpropagation in the Poincaré Fréchet mean graph convolution is $ {O}(L \cdot (n \cdot f^2 + T \cdot n \cdot d ))$, where $L$ denotes the number of layers, $f$ is the dimension of embedding features, $T$ is number of iterations for Fréchet mean convergence, and $d$ is average degree of the nodes. The complexity of the contrastive fusion module is ${O}(n \cdot f)$. The remainder no longer involves high computational complexity.

% ------------------- Methodology end --------------------------


% --------------------- Experiments ----------------------------
\section{Experiments}
\subsection{Dataset and Experimental Settings}
The Alzheimer’s Disease Neuroimaging Initiative (ADNI) \footnote{https://ida.loni.usc.edu/pages/access/studyData.jsp?project=ADNI} dataset is a comprehensive neuroimaging dataset for AD diagnosis. We evaluate 115 subjects, with 60 normal controls (NC), and 55 patients with AD for the experiments. Each participant has both DTI and fMRI data. 

To ensure the reproducibility of this study, we adopt QSIprep 0.19.1 and fMRIprep 20.2.3 to preprocess DTI and fMRI modalities respectively.
We analyze brain networks using the Automated Anatomical Labeling (AAL) Atlas, which contains 90 ROIs. Then the Pearson correlation coefficient and fractional anisotropy are used for fMRI and DTI brain graph construction. The hyperbolic graph convolutional model used in this work has two layers, with the curvature set to -1.0 and an embedding dimension of 128. During training, we apply a learning rate of 0.01 and the Adam optimizer to update network weights. To evaluate the model's performance, we conducted 5-fold cross-validation, repeating the experiments 10 times to ensure robust results.


\subsection{Baselines}
To establish a benchmark for comparison, we examined existing studies that leverage DTI and fMRI data for AD diagnosis. These studies incorporate advanced deep learning techniques, such as dynamic graph learning and hypergraph learning, to improve diagnostic accuracy. The state-of-the-art baseline methods are as follows:
\begin{itemize}
    \item[$-$] DecGAN \cite{pan2024decgan} employs a decoupling generative adversarial network to detect abnormal neural circuits associated with AD.
    \vspace{0.25em}
    \item[$-$] mmLasso \cite{meng2022multi} utilizes a multi-modal LassoNet framework to integrate supplementary information from different modalities. % DTI and fMRI
    \vspace{0.25em}
    \item[$-$] Cross-GNN \cite{yang2023mapping} captures inter-modal dependencies through dynamic graph learning and mutual learning. 
    \vspace{0.25em}
    \item[$-$] PALH \cite{zuo2024prior} predicts abnormal brain connections through a prior-guided adversarial learning framework with hypergraph. % DTI, fMRI, and PET


\end{itemize}


\subsection{Result Analysis}
We evaluate the performance of the model using accuracy (ACC), sensitivity (Sen.), and specificity (Spe.). The mean values achieved are \acc\% for accuracy, \sen\% for sensitivity, and \spe\% for specificity, with standard deviations (SD) of 1.10\%, 2.10\%, and 3.64\% respectively, as shown in Table \ref{tab_sota}. Since the baseline methods do not provide open-source code, their results were collected from their original papers. Our model achieved the highest accuracy and demonstrates strong overall classification performance and effectiveness. For example, while mmLasso \cite{meng2022multi} uses 16,830 dimensions for disease classification, our model only uses 128 dimensions, significantly reducing the feature space without compromising performance. Although our model's sensitivity and specificity were slightly below the best, it maintains a strong balance between correctly identifying true positives (sensitivity) and minimizing false positives (specificity), ensuring reliable disease detection. Overall, \algorithmName\ outperforms the baseline models in detecting Alzheimer's disease with improved accuracy and efficiency.


\begin{table}[ht]
    \centering
    \renewcommand{\arraystretch}{1.2}
    \begin{threeparttable}
    \caption{Classification performance comparison with baseline methods.} 

    \begin{tabular}{ m{2.8cm}   m{2.7cm} m{2.7cm} m{2.7cm} }
        \hline
        \textbf{Method} & \textbf{ACC(\%)}  & \textbf{Sen.(\%)}  & \textbf{Spe.(\%)}   \\
        \hline
        DecGAN \cite{pan2024decgan} &  85.18 & 81.25 & 90.91 \\
        mmLasso \cite{meng2022multi} &  90.68$\pm0.34$ & 88.81$\pm0.68$ & 91.91$\pm0.55$ \\
        Cross-GNN \cite{yang2023mapping} &  88.6$\pm6.3$ & \textbf{95.9$\pm9.2$} & 85.2$\pm7.7$ \\
        PALH \cite{zuo2024prior} &  88.73 & 84.37 & \textbf{92.31} \\

        \arrayrulecolor[HTML]{999999}\specialrule{0.05em}{0.05em}{0.05em} 
        \algorithmName & \textbf{\acc$\pm1.10$}  &  \sen$\pm2.10$  &  \spe$\pm3.64$ \\
        \arrayrulecolor{black}\hline
    \end{tabular}
    \label{tab_sota}
    \end{threeparttable}
\end{table}
% \vspace{-0.5em}


\subsection{Effectiveness Analysis of Hyperbolic Representation}
\subsubsection{Effect of Hyperbolic Curvature}
To evaluate the impact of hyperbolic curvature on the performance of our model, we conducted experiments across different curvature values, and curvature 0 means Euclidean space. This test is conducted with embedding dimension 16. 
The results are shown in Fig. \ref{fig:image2}(a). The hyperbolic and Euclidean results are colored blue and orange, respectively. Overall, our model with negative curvature outperforms the flat curvature in the classification task, suggesting that hyperbolic space representation is a superior choice for multimodal fusion in AD diagnosis.


\vspace{0.5em}
\begin{figure}[h]
    \begin{minipage}[b]{0.5\textwidth}
        \centering
        \begin{tikzpicture}
            \begin{axis}[
                ybar,
                bar width=.2cm,
                width=0.75\textwidth,
                height=0.65\textwidth,
                scale only axis,
                clip=false,
                separate axis lines,
                axis on top,
                xmin=0.5,
                xmax=6.5,
                xtick={1,2,3,4,5,6},
                xticklabels={-1.8,-1.4,-1.0,-0.6,-0.2,0},
                xlabel={(a) Effect of curvature},
                ytick={0,50,60,70, 80, 90,100},
                ymin=75,
                ymax=100,
                ylabel={ACC (\%)},
                ylabel style={at={(0.1,0.5)}},
                every axis plot/.append style={
                    ybar,
                    bar width=.35cm,
                    bar shift=0pt,
                },
                nodes near coords,
                nodes near coords align={vertical},
                every node near coord/.append style = {
                    yshift=0.2cm,
                    color=black,
                    anchor=south,
                    /pgf/number format/.cd,
                    fixed,
                    fixed zerofill,
                    precision=1
                },
            ]

            \addplot+[
                color = dorange,
                fill = dorange,
                pattern=north west lines,
                pattern color=dorange,
                error bars/.cd, 
                y dir=both,
                y explicit, 
            ]coordinates{(6,82.14) +- (0.0, 0.6)};
            \addplot+[
                color = dblue,
                fill = dblue,
                pattern=north east lines,
                pattern color=dblue,
                error bars/.cd, 
                y dir=both,
                y explicit, 
            ]coordinates{
                (1,92.86) +- (0.0, 0.5)
                (2,92.86) +- (0.0, 1.5)
                (3,94.05) +- (0.0, 1.5)
                (4,91.67) +- (0.0, 0.6)
                (5,90.48) +- (0.0, 0.9)
            };
            \end{axis}
        \end{tikzpicture}
    \end{minipage}
    \begin{minipage}[b]{0.5\textwidth}
        \centering

        \begin{tikzpicture}
            \begin{axis}[
                width=\textwidth,
                height=0.9\textwidth,
                xlabel={(b) Effect of embedding dimension},
                xmin=0.5,
                xmax=7.5,
                xtick={1,2,3,4,5,6,7},
                xticklabels={16,32,64,96,128,192,256},
                ylabel={ACC (\%)},
                ylabel style={at={(0.1,0.5)}},
                ymin=65,
                ymax=105,
                ytick={50,60,70,80,90,100},
                ybar=2*\pgflinewidth,
                bar width=6pt,
                scaled y ticks = false,
                legend entries={Euclidean,Hyperbolic},
                legend image code/.code={%
                    \draw[#1] (0cm,-0.1cm) rectangle (0.3cm,0.1cm);
                } 
                legend cell align=left,
                legend style={
                        at={(1,1.0)},
                        font=\fontsize{8}{8}\selectfont,
                        anchor=north east,
                },
                extra y ticks = 0.4,
                extra y tick labels={},
                extra y tick style={grid=major,major grid style={thick,draw=black}},
            ]

                \addplot+[
                    color = ddgreen,
                    fill = ddgreen,
                    pattern=north east lines,
                    pattern color=ddgreen,
                    error bars/.cd, 
                    y dir=both,
                    y explicit, 
                    % area legend
                ]
                    coordinates {
                        (1,82.14) +- (0.0, 1.1)
                        (2,79.00) +- (0.0, 1.0)
                        (3,84.52) +- (0.0, 1.2)
                        (4,83.33) +- (0.0, 0.8)
                        (5,82.14) +- (0.0, 0.5)
                        (6,77.38) +- (0.0, 1.1)
                        (7,72.62) +- (0.0, 1.2)
                    };

                \addplot+[
                    color = ddpurple,
                    fill = ddpurple,
                    pattern=north west lines,
                    pattern color=ddpurple,
                    error bars/.cd, 
                    y dir=both,
                    y explicit, 
                    % area legend
                ]
                     coordinates {
                        (1,95.24) +- (0.0, 0.7)
                        (2,95.24) +- (0.0, 1.2)
                        (3,92.86) +- (0.0, 0.5)
                        (4,94.05) +- (0.0, 0.7)
                        (5,92.86) +- (0.0, 1.5)
                        (6,92.86) +- (0.0, 1.1)
                        (7,91.67) +- (0.0, 0.5)
                     };
        
            \end{axis}
        \end{tikzpicture}

    \end{minipage}
    \caption{Performance comparison of different curvature values (a) and dimensions (b). In (a), the best result is observed at a curvature of -1.0, and worst is at curvature 0 which corresponds to Euclidean space. In (b), the ACC (\%) of the algorithm in Euclidean and hyperbolic space is compared.}
    \label{fig:image2}
\end{figure}


\subsubsection{Effect of Embedding Dimension}
We conducted experiments with different embedding dimensions and also reimplemented the \algorithmName\, using GCN in Euclidean space for comparison. The hyperbolic curvature is set to -0.8. The results are shown in Fig. \ref{fig:image2}(b). Our proposed algorithm achieves near-optimal performance with an embedding dimension of 16, which is significantly smaller than the 64-dimensional embedding required by GCN. This indicates that hyperbolic space embedding can significantly reduce the required embedding dimension while maintaining high performance.


\subsection{Ablation Study}
To evaluate the contributions of each component in the proposed model, we conducted an ablation study by sequentially replacing the hyperbolic space convolution with the Euclidean space convolution, removing the fusion modality, and both. The hyperbolic curvature is set to \curvature \, and embedding dimension is \dimension. The accuracy decreased by 11.42\%, 4.99\%, and 17.77\%, respectively, as shown in Table \ref{tab:ablation}. These results indicate that hyperbolic Poincaré Fréchet mean convolution significantly contributes to the overall performance of the model, also means proposed \algorithmName\, can greatly reduce the representation distortion compared to Euclidean space.


\begin{table}[ht]
    \centering
    \begin{threeparttable}
    \renewcommand{\arraystretch}{1.2}
    \caption{Contribution of each component.} 
    \vspace{0.5em}

    \begin{tabular}{ m{4.5cm}    m{1.6cm}  m{1.6cm}  m{1.6cm}  }
        \hline
        \textbf{Method}  & \textbf{ACC(\%)} & \textbf{Sen.(\%)} & \textbf{Spe.(\%)}  \\
        \hline
        \algorithmName  & \acc & \sen & \spe \\
        \arrayrulecolor[HTML]{999999}\specialrule{0.05em}{0.05em}{0.05em} 
        \algorithmName\,\textsubscript{wo:hyperbolic}  & 82.14 & 81.11 & 81.79 \\
        \algorithmName\,\textsubscript{wo:fusion} & 88.57 &	92.0 &	84.62    \\
        \algorithmName\,\textsubscript{wo:hyperbolic\&fusion}& 75.79 &	78.59 &	72.46     \\
        \arrayrulecolor{black}\hline
    \end{tabular}
    \begin{tablenotes}
        \small
        \item 'wo' indicates the removal of a module from the \algorithmName.
    \end{tablenotes}

    \label{tab:ablation}
    \end{threeparttable}
\end{table}
% --------------------- Experiments end ----------------------------


% --------------------- Conclusion ----------------------------
\section{Conclusion}
In this paper, we have proposed a novel multimodal hyperbolic graph convolutional neural network for brain network analysis. The proposed method leverages the hyperbolic space to model the intrinsic geometry of the brain network. We conducted experiments on ADNI dataset and demonstrated that the proposed method outperforms baseline methods. The results show that the proposed method achieves a \acc\% classification accuracy with 115 ADNI subjects. This performance not only outperforms baseline methods but also underscores the efficacy of our approach in handling complex neuroimaging data. We also conducted an ablation study to investigate the effect of hyperbolic curvature, embedding dimension, and fusion methods on the performance of the model. The proposed model offers a new perspective in brain network analysis, with significant implications for the diagnosis and treatment of brain disorders. By enhancing the accuracy and efficacy of neuroimaging analyses, our approach holds the potential to greatly improve clinical outcomes for patients with Alzheimer's disease and other neurodegenerative disorders.
% --------------------- Conclusion end ----------------------------


% ---------------------- Bibliography -----------------------
\bibliographystyle{splncs04}
\bibliography{references.bib}

% ---------------------- Bibliography end  -------------------------

\end{document}
