## Supplementary material for "Multimodal Hyperbolic Graph Learning for Alzheimer’s Disease Detection": latex: references.bib

%%%%%%%%%%%%%%%%%% Alzheimer's   disease
@article{scheltens2021alzheimer,
  title={Alzheimer's disease},
  author={Scheltens, Philip and De Strooper, Bart and Kivipelto, Miia and Holstege, Henne and Ch{\'e}telat, Gael and Teunissen, Charlotte E and Cummings, Jeffrey and van der Flier, Wiesje M},
  journal={The Lancet},
  volume={397},
  number={10284},
  pages={1577--1590},
  year={2021},
  publisher={Elsevier}
}

@article{selkoe2024advent,
  title={The advent of Alzheimer treatments will change the trajectory of human aging},
  author={Selkoe, Dennis J},
  journal={Nature Aging},
  volume={4},
  number={4},
  pages={453--463},
  year={2024},
  publisher={Nature Publishing Group US New York}
}

@article{nandi2024cost,
  title={Cost of care for Alzheimer’s disease and related dementias in the United States: 2016 to 2060},
  author={Nandi, Arindam and Counts, Nathaniel and Br{\"o}ker, Janina and Malik, Sabrina and Chen, Simiao and Han, Rachael and Klusty, Jessica and Seligman, Benjamin and Tortorice, Daniel and Vigo, Daniel and others},
  journal={npj Aging},
  volume={10},
  number={1},
  pages={13},
  year={2024},
  publisher={Nature Publishing Group UK London}
}

@ARTICLE{Gustavsson2023-ki,
  title    = "Global estimates on the number of persons across the Alzheimer's
              disease continuum",
  author   = "Gustavsson, Anders and Norton, Nicholas and Fast, Thomas and
              Fr{\"o}lich, Lutz and Georges, Jean and Holzapfel, Drew and
              Kirabali, Tunahan and Krolak-Salmon, Pierre and Rossini, Paolo M
              and Ferretti, Maria Teresa and Lanman, Lydia and Chadha,
              Antonella Santuccione and van der Flier, Wiesje M",
  abstract = "INTRODUCTION: Global estimates on numbers of persons in early
              stages of Alzheimer's disease (AD), including prodromal and
              preclinical, are lacking, yet are needed to inform policy
              decisions on preventive measures and planning for future
              therapies targeting AD pathology. METHODS: We synthesized the
              literature on prevalence across the AD continuum and derived a
              model estimating the number of persons, stratified by 5-year age
              groups, sex, and disease stage (AD dementia, prodromal AD, and
              preclinical AD). RESULTS: The global number of persons with AD
              dementia, prodromal AD, and preclinical AD were estimated at 32,
              69, and 315 million, respectively. Together they constituted 416
              million across the AD continuum, or 22\% of all persons aged 50
              and above. DISCUSSION: Considering predementia stages, the number
              of persons with AD is much larger than conveyed in available
              literature. Our estimates are uncertain, especially for
              predementia stages in low- and middle-income regions where
              biomarker studies are missing.",
  journal  = "Alzheimers. Dement.",
  volume   =  19,
  number   =  2,
  pages    = "658--670",
  month_fake    =  feb,
  year     =  2023,
  keywords = "Alzheimer's disease; preclinical; prevalence; prevention;
              prodromal",
  language = "en"
}

@ARTICLE{Nandi2022-fb,
  title     = "Global and regional projections of the economic burden of
               Alzheimer's disease and related dementias from 2019 to 2050: A
               value of statistical life approach",
  author    = "Nandi, Arindam and Counts, Nathaniel and Chen, Simiao and
               Seligman, Benjamin and Tortorice, Daniel and Vigo, Daniel and
               Bloom, David E",
  abstract  = "Background: The burden of Alzheimer's disease and related
               dementias (ADRDs) is expected to grow rapidly with population
               aging, especially in low- and middle-income countries, in the
               next few decades. We used a willingness-to-pay approach to
               project the global, regional, and national economic burden of
               ADRDs from 2019 to 2050 under status quo. Methods: We projected
               age group and country-specific disability-adjusted life years
               (DALYs) lost to ADRDs in future years based on historical growth
               in disease burden and available population projections. We used
               country-specific extrapolations of the value of a statistical
               life (VSL) year and its future projections based on historical
               income growth to estimate the economic burden - measured in
               terms of the value of lost DALYs - of ADRDs. A probabilistic
               uncertainty analysis was used to calculate point estimates and
               95\% uncertainty bounds of the economic burden. Findings: In
               2019, the global VSL-based economic burden of ADRDs was an
               estimated $2.8 trillion. The burden was projected to increase to
               $4.7 trillion (95\% uncertainty bound: $4 trillion-$5.5
               trillion) in 2030, $8.5 trillion ($6.8 trillion-$10.8 trillion)
               in 2040, and $16.9 trillion ($11.3 trillion-$27.3 trillion) in
               2050. Low- and middle-income countries (LMICs) would account for
               65\% of the global VSL-based economic burden in 2050, as
               compared with only 18\% in 2019. Within LMICs, upper-middle
               income countries would carry the largest VSL-based economic
               burden by 2050 (92\% of LMICs burden and 60\% of global burden).
               Interpretation: ADRDs have a large and inequitable projected
               future VSL-based economic burden. Funding: The Davos Alzheimer's
               Collaborative.",
  journal   = "EClinicalMedicine",
  publisher = "Elsevier BV",
  volume    =  51,
  number    =  101580,
  pages     = "101580",
  month_fake     =  sep,
  year      =  2022,
  keywords  = "ADRD; Alzheimer's disease and related dementias; Dementia; VSL;
               Value of statistical life",
  copyright = "http://creativecommons.org/licenses/by-nc-nd/4.0/",
  language  = "en"
}


%%%%%%%%%%%%%%%%%% graph learning
@article{Xia2021,
  title = {Graph Learning: A Survey},
  volume = {2},
  ISSN = {2691-4581},
  number = {2},
  journal = {IEEE Transactions on Artificial Intelligence},
  publisher = {Institute of Electrical and Electronics Engineers (IEEE)},
  author = {Xia,  Feng and Sun,  Ke and Yu,  Shuo and Aziz,  Abdul and Wan,  Liangtian and Pan,  Shirui and Liu,  Huan},
  year = {2021},
  month_fake = apr,
  pages = {109–127}
}


@ARTICLE{Zhou2020-vy,
  title     = "Graph neural networks: A review of methods and applications",
  author    = "Zhou, Jie and Cui, Ganqu and Hu, Shengding and Zhang, Zhengyan
               and Yang, Cheng and Liu, Zhiyuan and Wang, Lifeng and Li,
               Changcheng and Sun, Maosong",
  journal   = "AI Open",
  publisher = "Elsevier BV",
  volume    =  1,
  pages     = "57--81",
  year      =  2020,
  copyright = "http://creativecommons.org/licenses/by/4.0/",
  language  = "en"
}

@ARTICLE{Wu2021-ut,
  title     = "A comprehensive survey on graph neural networks",
  author    = "Wu, Zonghan and Pan, Shirui and Chen, Fengwen and Long, Guodong
               and Zhang, Chengqi and Yu, Philip S",
  abstract  = "Deep learning has revolutionized many machine learning tasks in
               recent years, ranging from image classification and video
               processing to speech recognition and natural language
               understanding. The data in these tasks are typically represented
               in the Euclidean space. However, there is an increasing number
               of applications, where data are generated from non-Euclidean
               domains and are represented as graphs with complex relationships
               and interdependency between objects. The complexity of graph
               data has imposed significant challenges on the existing machine
               learning algorithms. Recently, many studies on extending deep
               learning approaches for graph data have emerged. In this
               article, we provide a comprehensive overview of graph neural
               networks (GNNs) in data mining and machine learning fields. We
               propose a new taxonomy to divide the state-of-the-art GNNs into
               four categories, namely, recurrent GNNs, convolutional GNNs,
               graph autoencoders, and spatial-temporal GNNs. We further
               discuss the applications of GNNs across various domains and
               summarize the open-source codes, benchmark data sets, and model
               evaluation of GNNs. Finally, we propose potential research
               directions in this rapidly growing field.",
  journal   = "IEEE Trans. Neural Netw. Learn. Syst.",
  publisher = "Institute of Electrical and Electronics Engineers (IEEE)",
  volume    =  32,
  number    =  1,
  pages     = "4--24",
  month_fake     =  jan,
  year      =  2021,
  copyright = "https://ieeexplore.ieee.org/Xplorehelp/downloads/license-information/IEEE.html",
  language  = "en"
}


%%%%%%%%%%%%%%%%%% graph learning for AD
@ARTICLE{Chen2023-bi,
  title     = "Orthogonal latent space learning with feature weighting and
               graph learning for multimodal Alzheimer's disease diagnosis",
  author    = "Chen, Zhi and Liu, Yongguo and Zhang, Yun and Li, Qiaoqin and
               {Alzheimer's Disease Neuroimaging Initiative}",
  abstract  = "Recent studies have shown that multimodal neuroimaging data
               provide complementary information of the brain and latent
               space-based methods have achieved promising results in fusing
               multimodal data for Alzheimer's disease (AD) diagnosis. However,
               most existing methods treat all features equally and adopt
               nonorthogonal projections to learn the latent space, which
               cannot retain enough discriminative information in the latent
               space. Besides, they usually preserve the relationships among
               subjects in the latent space based on the similarity graph
               constructed on original features for performance boosting.
               However, the noises and redundant features significantly corrupt
               the graph. To address these limitations, we propose an
               Orthogonal Latent space learning with Feature weighting and
               Graph learning (OLFG) model for multimodal AD diagnosis.
               Specifically, we map multiple modalities into a common latent
               space by orthogonal constrained projection to capture the
               discriminative information for AD diagnosis. Then, a feature
               weighting matrix is utilized to sort the importance of features
               in AD diagnosis adaptively. Besides, we devise a regularization
               term with learned graph to preserve the local structure of the
               data in the latent space and integrate the graph construction
               into the learning processing for accurately encoding the
               relationships among samples. Instead of constructing a
               similarity graph for each modality, we learn a joint graph for
               multiple modalities to capture the correlations among
               modalities. Finally, the representations in the latent space are
               projected into the target space to perform AD diagnosis. An
               alternating optimization algorithm with proved convergence is
               developed to solve the optimization objective. Extensive
               experimental results show the effectiveness of the proposed
               method.",
  journal   = "Med. Image Anal.",
  publisher = "Elsevier BV",
  volume    =  84,
  number    =  102698,
  pages     = "102698",
  month_fake     =  feb,
  year      =  2023,
  keywords  = "Alzheimer's disease; Graph learning; Latent representation
               learning; Mild cognitive impairment; Multimodal fusion",
  language  = "en"
}


@ARTICLE{Zhang2023-yw, 
  title     = "Multi-modal graph neural network for early diagnosis of
               Alzheimer's disease from {sMRI} and {PET} scans",
  author    = "Zhang, Yanteng and He, Xiaohai and Chan, Yi Hao and Teng, Qizhi
               and Rajapakse, Jagath C",
  abstract  = "In recent years, deep learning models have been applied to
               neuroimaging data for early diagnosis of Alzheimer's disease
               (AD). Structural magnetic resonance imaging (sMRI) and positron
               emission tomography (PET) images provide structural and
               functional information about the brain, respectively. Combining
               these features leads to improved performance than using a single
               modality alone in building predictive models for AD diagnosis.
               However, current multi-modal approaches in deep learning, based
               on sMRI and PET, are mostly limited to convolutional neural
               networks, which do not facilitate integration of both image and
               phenotypic information of subjects. We propose to use graph
               neural networks (GNN) that are designed to deal with problems in
               non-Euclidean domains. In this study, we demonstrate how brain
               networks are created from sMRI or PET images and can be used in
               a population graph framework that combines phenotypic
               information with imaging features of the brain networks. Then,
               we present a multi-modal GNN framework where each modality has
               its own branch of GNN and a technique that combines the
               multi-modal data at both the level of node vectors and adjacency
               matrices. Finally, we perform late fusion to combine the
               preliminary decisions made in each branch and produce a final
               prediction. As multi-modality data becomes available,
               multi-source and multi-modal is the trend of AD diagnosis. We
               conducted explorative experiments based on multi-modal imaging
               data combined with non-imaging phenotypic information for AD
               diagnosis and analyzed the impact of phenotypic information on
               diagnostic performance. Results from experiments demonstrated
               that our proposed multi-modal approach improves performance for
               AD diagnosis. Our study also provides technical reference and
               support the need for multivariate multi-modal diagnosis methods.",
  journal   = "Comput. Biol. Med.",
  publisher = "Elsevier BV",
  volume    =  164,
  number    =  107328,
  pages     = "107328",
  month_fake     =  sep,
  year      =  2023,
  keywords  = "Alzheimer's disease diagnosis; Brain networks; Graph neural
               networks; Multi-modal method; PET; sMRI",
  language  = "en"
}


@ARTICLE{Qu2023-xv,
  title     = "A graph convolutional network based on univariate
               neurodegeneration biomarker for Alzheimer's disease diagnosis",
  author    = "Qu, Zongshuai and Yao, Tao and Liu, Xinghui and Wang, Gang",
  abstract  = "OBJECTIVE: Alzheimer's disease (AD) is a progressive and
               irreversible neurodegenerative disease that is not easily
               detectable in the early stage. This study proposed an efficient
               method of applying a graph convolutional network (GCN) on the
               early prediction of AD. METHODS: We proposed a univariate
               neurodegeneration biomarker (UNB) based GCN semi-supervised
               classification framework. We generated UNB by comparing the
               similarity of individual morphological atrophy pattern and the
               atrophy pattern of [Formula: see text] AD group according to the
               brain morphological abnormalities induced by AD. For the GCN
               semi-supervised classification model, we took the UNBs of
               individuals as the features of nodes and constructed the weight
               of edges according to the similarity of phenotypic information
               between individuals, which explored the essential features of
               individuals through spectral graph convolution. The attention
               module was constructed and embedded into the GCN framework,
               which may refine the input morphological features to highlight
               the main impact of AD on the cerebral cortex and weaken the
               instability caused by individual diversities, thereby
               identifying the significant ROIs affected by AD and improving
               the classification accuracy. RESULTS: We tested the UNB-GCN
               framework on the Alzheimer's Disease Neuroimaging Initiative
               (ADNI) database. The estimated minimum sample sizes were 156,
               349 and 423 for the longitudinal [Formula: see text] AD,
               [Formula: see text] mild cognitive impairment (MCI) and
               [Formula: see text] cognitively unimpaired (CU) groups,
               respectively. And the proposed UNB-GCN framework combined with
               the attention module can effectively improve the classification
               performance with 93.90\% classification accuracy for AD vs. CU
               and 82.05\% for AD vs. MCI on the validation set. CONCLUSION:
               The proposed UNB measures were superior to the conventional
               volume measures in describing the AD-induced cerebral cortex
               morphological changes. And the UNB-GCN framework combined with
               attention module may effectively improve the classification
               performance between MCI subjects and AD patients. Clinical and
               Translational Impact Statement: This study aims to predict the
               early AD patients, so as to help clinicians develop effective
               interventions to delay the deterioration of AD symptoms.",
  journal   = "IEEE J. Transl. Eng. Health Med.",
  publisher = "Institute of Electrical and Electronics Engineers (IEEE)",
  volume    =  11,
  pages     = "405--416",
  month_fake     =  jun,
  year      =  2023,
  keywords  = "Alzheimer's disease; attention module; graph convolutional
               network; magnetic resonance imaging; univariate
               neurodegeneration biomarker",
  copyright = "https://creativecommons.org/licenses/by/4.0/legalcode",
  language  = "en"
}

@ARTICLE{Shan2022-la,
  title     = "Spatial-temporal graph convolutional network for Alzheimer
               classification based on brain functional connectivity imaging of
               electroencephalogram",
  author    = "Shan, Xiaocai and Cao, Jun and Huo, Shoudong and Chen, Liangyu
               and Sarrigiannis, Ptolemaios Georgios and Zhao, Yifan",
  abstract  = "Functional connectivity of the human brain, representing
               statistical dependence of information flow between cortical
               regions, significantly contributes to the study of the intrinsic
               brain network and its functional mechanism. To fully explore its
               potential in the early diagnosis of Alzheimer's disease (AD)
               using electroencephalogram (EEG) recordings, this article
               introduces a novel dynamical spatial-temporal graph
               convolutional neural network (ST-GCN) for better classification
               performance. Different from existing studies that are based on
               either topological brain function characteristics or temporal
               features of EEG, the proposed ST-GCN considers both the
               adjacency matrix of functional connectivity from multiple EEG
               channels and corresponding dynamics of signal EEG channel
               simultaneously. Different from the traditional graph
               convolutional neural networks, the proposed ST-GCN makes full
               use of the constrained spatial topology of functional
               connectivity and the discriminative dynamic temporal information
               represented by the 1D convolution. We conducted extensive
               experiments on the clinical EEG data set of AD patients and
               Healthy Controls. The results demonstrate that the proposed
               method achieves better classification performance (92.3\%) than
               the state-of-the-art methods. This approach can not only help
               diagnose AD but also better understand the effect of normal
               ageing on brain network characteristics before we can accurately
               diagnose the condition based on resting-state EEG.",
  journal   = "Hum. Brain Mapp.",
  publisher = "Wiley",
  volume    =  43,
  number    =  17,
  pages     = "5194--5209",
  month_fake     =  dec,
  year      =  2022,
  keywords  = "artificial intelligence; brain association;
               electroencephalogram; graph convolutional neural network;
               machine learning",
  copyright = "http://creativecommons.org/licenses/by/4.0/",
  language  = "en"
}

@ARTICLE{Gao2023-kr,
  title     = "Brain age prediction using the graph neural network based on
               resting-state functional {MRI} in Alzheimer's disease",
  author    = "Gao, Jingjing and Liu, Jiaxin and Xu, Yuhang and Peng, Dawei and
               Wang, Zhengning",
  abstract  = "Introduction: Alzheimer's disease (AD) is a neurodegenerative
               disease that significantly impacts the quality of life of
               patients and their families. Neuroimaging-driven brain age
               prediction has been proposed as a potential biomarker to detect
               mental disorders, such as AD, aiding in studying its effects on
               functional brain networks. Previous studies have shown that
               individuals with AD display impaired resting-state functional
               connections. However, most studies on brain age prediction have
               used structural magnetic resonance imaging (MRI), with limited
               studies based on resting-state functional MRI (rs-fMRI).
               Methods: In this study, we applied a graph neural network (GNN)
               model on controls to predict brain ages using rs-fMRI in
               patients with AD. We compared the performance of the GNN model
               with traditional machine learning models. Finally, the post hoc
               model was also used to identify the critical brain regions in
               AD. Results: The experimental results demonstrate that our GNN
               model can predict brain ages of normal controls using rs-fMRI
               data from the ADNI database. Moreover the differences between
               brain ages and chronological ages were more significant in AD
               patients than in normal controls. Our results also suggest that
               AD is associated with accelerated brain aging and that the GNN
               model based on resting-state functional connectivity is an
               effective tool for predicting brain age. Discussion: Our study
               provides evidence that rs-fMRI is a promising modality for brain
               age prediction in AD research, and the GNN model proves to be
               effective in predicting brain age. Furthermore, the effects of
               the hippocampus, parahippocampal gyrus, and amygdala on brain
               age prediction are verified.",
  journal   = "Front. Neurosci.",
  publisher = "Frontiers Media SA",
  volume    =  17,
  pages     = "1222751",
  month_fake     =  jun,
  year      =  2023,
  keywords  = "Alzheimer's disease; age prediction; brain aging; graph neural
               network; resting-state functional magnetic resonance imaging",
  copyright = "https://creativecommons.org/licenses/by/4.0/",
  language  = "en"
}

@ARTICLE{Zhu2022-vw,
  title     = "Interpretable learning based Dynamic Graph Convolutional
               Networks for Alzheimer's Disease analysis",
  author    = "Zhu, Yonghua and Ma, Junbo and Yuan, Changan and Zhu, Xiaofeng",
  journal   = "Inf. Fusion",
  publisher = "Elsevier BV",
  volume    =  77,
  pages     = "53--61",
  month_fake     =  jan,
  year      =  2022,
  language  = "en"
}


@INPROCEEDINGS{Peng2024-ws,
  title           = "Stage-aware Brain Graph Learning for Alzheimer's Disease",
  booktitle       = "The 2024 IEEE Conference on Artificial Intelligence (CAI)",
  author          = "Peng, Ciyuan and Liu, Mujie and Meng, Chenxuan and Xue, Sha and Keogh, Kathleen and Xia, Feng",
  month_fake           =  jun,
  year            =  2024,
  conference      = "The 2024 IEEE Conference on Artificial Intelligence (CAI)",
  location        = "Singapore"
}

@ARTICLE{Zeng2022-kn,
  title     = "Graph convolutional network with sample and feature weights for
               Alzheimer's disease diagnosis",
  author    = "Zeng, Lu and Li, Hengxin and Xiao, Tingsong and Shen, Fumin and
               Zhong, Zhi",
  journal   = "Inf. Process. Manag.",
  publisher = "Elsevier BV",
  volume    =  59,
  number    =  4,
  pages     = "102952",
  month_fake     =  jul,
  year      =  2022,
  language  = "en"
}

@ARTICLE{Zhang2023-el,
  title     = "Multi-relation graph convolutional network for Alzheimer's
               disease diagnosis using structural {MRI}",
  author    = "Zhang, Jin and He, Xiaohai and Qing, Linbo and Chen, Xiang and
               Liu, Yan and Chen, Honggang",
  journal   = "Knowl. Based Syst.",
  publisher = "Elsevier BV",
  volume    =  270,
  number    =  110546,
  pages     = "110546",
  month_fake     =  jun,
  year      =  2023,
  language  = "en"
}

@ARTICLE{De_Mendonca2023-sh,
  title     = "Alzheimer's disease classification based on graph kernel {SVMs}
               constructed with {3D} texture features extracted from {MR}
               images",
  author    = "de Mendon{\c c}a, Lucas Jos{\'e} Cruz and Ferrari, Ricardo
               Jos{\'e}",
  journal   = "Expert Syst. Appl.",
  publisher = "Elsevier BV",
  volume    =  211,
  number    =  118633,
  pages     = "118633",
  month_fake     =  jan,
  year      =  2023,
  language  = "en"
}

@ARTICLE{Xu2024-jc,
  title     = "Interpretable medical deep framework by logits-constraint
               attention guiding graph-based multi-scale fusion for Alzheimer's
               disease analysis",
  author    = "Xu, Jinghao and Yuan, Chenxi and Ma, Xiaochuan and Shang,
               Huifang and Shi, Xiaoshuang and Zhu, Xiaofeng",
  journal   = "Pattern Recognit.",
  publisher = "Elsevier BV",
  volume    =  152,
  number    =  110450,
  pages     = "110450",
  month_fake     =  aug,
  year      =  2024,
  language  = "en"
}

@ARTICLE{Hett2021-lh,
  title     = "Multi-scale graph-based grading for Alzheimer's disease
               prediction",
  author    = "Hett, Kilian and Ta, Vinh-Thong and Oguz, Ipek and Manj{\'o}n,
               Jos{\'e} V and Coup{\'e}, Pierrick and {Alzheimer's Disease
               Neuroimaging Initiative}",
  abstract  = "The prediction of subjects with mild cognitive impairment (MCI)
               who will progress to Alzheimer's disease (AD) is clinically
               relevant, and may above all have a significant impact on
               accelerating the development of new treatments. In this paper,
               we present a new MRI-based biomarker that enables us to
               accurately predict conversion of MCI subjects to AD. In order to
               better capture the AD signature, we introduce two main
               contributions. First, we present a new graph-based grading
               framework to combine inter-subject similarity features and
               intra-subject variability features. This framework involves
               patch-based grading of anatomical structures and graph-based
               modeling of structure alteration relationships. Second, we
               propose an innovative multiscale brain analysis to capture
               alterations caused by AD at different anatomical levels. Based
               on a cascade of classifiers, this multiscale approach enables
               the analysis of alterations of whole brain structures and
               hippocampus subfields at the same time. During our experiments
               using the ADNI-1 dataset, the proposed multiscale graph-based
               grading method obtained an area under the curve (AUC) of 81\% to
               predict conversion of MCI subjects to AD within three years.
               Moreover, when combined with cognitive scores, the proposed
               method obtained 85\% of AUC. These results are competitive in
               comparison to state-of-the-art methods evaluated on the same
               dataset.",
  journal   = "Med. Image Anal.",
  publisher = "Elsevier BV",
  volume    =  67,
  number    =  101850,
  pages     = "101850",
  month_fake     =  jan,
  year      =  2021,
  keywords  = "Alzheimer's disease classification; Graph-based method;
               Hippocampal subfields; Inter-subject similarity; Intra-subject
               variability; Mild cognitive impairment; Patch-based grading;
               Whole brain analysis",
  language  = "en"
}


%%%%%%%%%%%%%%%%%% multimodal graph learning for brain
@ARTICLE{Xu2021-dy,
  title     = "The integrated understanding of structural and functional
               connectomes in depression: A multimodal meta-analysis of graph
               metrics",
  author    = "Xu, Shu-Xian and Deng, Wen-Feng and Qu, Ying-Ying and Lai,
               Wen-Tao and Huang, Tan-Yu and Rong, Han and Xie, Xin-Hui",
  abstract  = "BACKGROUND: From the perspective of information processing, an
               integrated understanding of the structural and functional
               connectomes in depression patients is important, a multimodal
               meta-analysis is required to detect the robust alterations in
               graph metrics across studies. METHODS: Following a systematic
               search, 952 depression patients and 1447 controls in nine
               diffusion magnetic resonance imaging (dMRI) and twelve rest
               state functional MRI (rs-fMRI) studies with high methodological
               quality met the inclusion criteria and were included in the
               meta-analysis. RESULTS: Regarding the dMRI results, no
               significant differences of meta-analytic metrics were found;
               regarding the rs-fMRI results, the modularity and local
               efficiency were found to be significantly lower in the
               depression group than in the controls (Hedge's g = -0.330 and
               -0.349, respectively). CONCLUSION: Our findings suggested a
               lower modularity and network efficiency in the rs-fMRI network
               in depression patients, indicating that the pathological
               imbalances in brain connectomes needs further exploration.
               LIMITATIONS: Included number of trials was low and heterogeneity
               should be noted.",
  journal   = "J. Affect. Disord.",
  publisher = "Elsevier BV",
  volume    =  295,
  pages     = "759--770",
  month_fake     =  dec,
  year      =  2021,
  keywords  = "Brain connectome; Depression; Diffusion MRI; Graph theory;
               Meta-analysis; Rest state functional MRI",
  language  = "en"
}

@ARTICLE{Chen2023-rr,
  title     = "Discriminative analysis of schizophrenia patients using graph
               convolutional networks: A combined multimodal {MRI} and
               connectomics analysis",
  author    = "Chen, Xiaoyi and Ke, Pengfei and Huang, Yuanyuan and Zhou, Jing
               and Li, Hehua and Peng, Runlin and Huang, Jiayuan and Liang,
               Liqing and Ma, Guolin and Li, Xiaobo and Ning, Yuping and Wu,
               Fengchun and Wu, Kai",
  abstract  = "Introduction: Recent studies in human brain connectomics with
               multimodal magnetic resonance imaging (MRI) data have widely
               reported abnormalities in brain structure, function and
               connectivity associated with schizophrenia (SZ). However, most
               previous discriminative studies of SZ patients were based on MRI
               features of brain regions, ignoring the complex relationships
               within brain networks. Methods: We applied a graph convolutional
               network (GCN) to discriminating SZ patients using the features
               of brain region and connectivity derived from a combined
               multimodal MRI and connectomics analysis. Structural magnetic
               resonance imaging (sMRI) and resting-state functional magnetic
               resonance imaging (rs-fMRI) data were acquired from 140 SZ
               patients and 205 normal controls. Eighteen types of brain graphs
               were constructed for each subject using 3 types of node
               features, 3 types of edge features, and 2 brain atlases. We
               investigated the performance of 18 brain graphs and used the
               TopK pooling layers to highlight salient brain regions (nodes in
               the graph). Results: The GCN model, which used functional
               connectivity as edge features and multimodal features (sMRI +
               fMRI) of brain regions as node features, obtained the highest
               average accuracy of 95.8\%, and outperformed other existing
               classification studies in SZ patients. In the explainability
               analysis, we reported that the top 10 salient brain regions,
               predominantly distributed in the prefrontal and occipital
               cortices, were mainly involved in the systems of emotion and
               visual processing. Discussion: Our findings demonstrated that
               GCN with a combined multimodal MRI and connectomics analysis can
               effectively improve the classification of SZ at an individual
               level, indicating a promising direction for the diagnosis of SZ
               patients. The code is available at
               https://github.com/CXY-scut/GCN-SZ.git.",
  journal   = "Front. Neurosci.",
  publisher = "Frontiers Media SA",
  volume    =  17,
  pages     = "1140801",
  month_fake     =  mar,
  year      =  2023,
  keywords  = "discriminative analysis; graph convolutional network; human
               brain connectomics; multimodal MRI; schizophrenia",
  copyright = "https://creativecommons.org/licenses/by/4.0/",
  language  = "en"
}

@ARTICLE{Chen2023-ox,
  title    = "Corrigendum: Discriminative analysis of schizophrenia patients
              using graph convolutional networks: a combined multimodal {MRI}
              and connectomics analysis",
  author   = "Chen, Xiaoyi and Ke, Pengfei and Huang, Yuanyuan and Zhou, Jing
              and Li, Hehua and Peng, Runlin and Huang, Jiayuan and Liang,
              Liqin and Ma, Guolin and Li, Xiaobo and Ning, Yuping and Wu,
              Fengchun and Wu, Kai",
  abstract = "[This corrects the article DOI: 10.3389/fnins.2023.1140801.].",
  journal  = "Front. Neurosci.",
  volume   =  17,
  pages    = "1228322",
  month_fake    =  jun,
  year     =  2023,
  keywords = "discriminative analysis; graph convolutional network; human brain
              connectomics; multimodal MRI; schizophrenia",
  language = "en"
}

@INPROCEEDINGS{Zhou2022-lz,
  title           = "Interpretable graph convolutional network of
                     multi-modality brain imaging for Alzheimer's disease
                     diagnosis",
  booktitle       = "2022 {IEEE} 19th International Symposium on Biomedical
                     Imaging ({ISBI})",
  author          = "Zhou, Houliang and He, Lifang and Zhang, Yu and Shen, Li
                     and Chen, Brian",
  publisher       = "IEEE",
  month_fake           =  mar,
  year            =  2022,
  conference      = "2022 IEEE 19th International Symposium on Biomedical
                     Imaging (ISBI)",
  location        = "Kolkata, India"
}

@ARTICLE{Bi2023-vg,
  title    = "Community graph convolution neural network for Alzheimer's
              disease classification and pathogenetic factors identification",
  author   = "Bi, Xia-An and Chen, Ke and Jiang, Siyu and Luo, Sheng and Zhou,
              Wenyan and Xing, Zhaoxu and Xu, Luyun and Liu, Zhengliang and
              Liu, Tianming",
  abstract = "As a complex neural network system, the brain regions and genes
              collaborate to effectively store and transmit information. We
              abstract the collaboration correlations as the brain region gene
              community network (BG-CN) and present a new deep learning
              approach, such as the community graph convolutional neural
              network (Com-GCN), for investigating the transmission of
              information within and between communities. The results can be
              used for diagnosing and extracting causal factors for Alzheimer's
              disease (AD). First, an affinity aggregation model for BG-CN is
              developed to describe intercommunity and intracommunity
              information transmission. Second, we design the Com-GCN
              architecture with intercommunity convolution and intracommunity
              convolution operations based on the affinity aggregation model.
              Through sufficient experimental validation on the AD neuroimaging
              initiative (ADNI) dataset, the design of Com-GCN matches the
              physiological mechanism better and improves the interpretability
              and classification performance. Furthermore, Com-GCN can identify
              lesioned brain regions and disease-causing genes, which may
              assist precision medicine and drug design in AD and serve as a
              valuable reference for other neurological disorders.",
  journal  = "IEEE Trans. Neural Netw. Learn. Syst.",
  volume   = "PP",
  month_fake    =  may,
  year     =  2023,
  language = "en"
}

@ARTICLE{Song2021-nd,
  title     = "Auto-metric graph neural network based on a meta-learning
               strategy for the diagnosis of Alzheimer's disease",
  author    = "Song, Xiaofan and Mao, Mingyi and Qian, Xiaohua",
  abstract  = "Alzheimer's disease (AD) is the most common cognitive disorder.
               In recent years, many computer-aided diagnosis techniques have
               been proposed for AD diagnosis and progression predictions.
               Among them, graph neural networks (GNNs) have received extensive
               attention owing to their ability to effectively fuse multimodal
               features and model the correlation between samples. However,
               many GNNs for node classification use an entire dataset to
               construct a large fixed-graph structure, which cannot be used
               for independent testing. To overcome this limitation while
               maintaining the advantages of the GNN, we propose an auto-metric
               GNN (AMGNN) model for AD diagnosis. First, a metric-based
               meta-learning strategy is introduced to realize inductive
               learning for independent testing through multiple node
               classification tasks. In the meta-tasks, the small graphs help
               make the model insensitive to the sample size, thus improving
               the performance under small sample size conditions. Furthermore,
               an AMGNN layer with a probability constraint is designed to
               realize node similarity metric learning and effectively fuse
               multimodal data. We verified the model on two tasks based on the
               TADPOLE dataset: early AD diagnosis and mild cognitive
               impairment (MCI) conversion prediction. Our model provides
               excellent performance on both tasks with accuracies of 94.44\%
               and 87.50\% and median accuracies of 94.19\% and 86.25\%,
               respectively. These results show that our model improves
               flexibility while ensuring a good classification performance,
               thus promoting the development of graph-based deep learning
               algorithms for disease diagnosis.",
  journal   = "IEEE J. Biomed. Health Inform.",
  publisher = "Institute of Electrical and Electronics Engineers (IEEE)",
  volume    =  25,
  number    =  8,
  pages     = "3141--3152",
  month_fake     =  aug,
  year      =  2021,
  copyright = "https://ieeexplore.ieee.org/Xplorehelp/downloads/license-information/IEEE.html",
  language  = "en"
}

@ARTICLE{Zhang2022-uh,
  title     = "Graph-based autoencoder integrates spatial transcriptomics with
               chromatin images and identifies joint biomarkers for Alzheimer's
               disease",
  author    = "Zhang, Xinyi and Wang, Xiao and Shivashankar, G V and Uhler,
               Caroline",
  abstract  = "Tissue development and disease lead to changes in cellular
               organization, nuclear morphology, and gene expression, which can
               be jointly measured by spatial transcriptomic technologies.
               However, methods for jointly analyzing the different spatial
               data modalities in 3D are still lacking. We present a
               computational framework to integrate Spatial Transcriptomic data
               using over-parameterized graph-based Autoencoders with Chromatin
               Imaging data (STACI) to identify molecular and functional
               alterations in tissues. STACI incorporates multiple modalities
               in a single representation for downstream tasks, enables the
               prediction of spatial transcriptomic data from nuclear images in
               unseen tissue sections, and provides built-in batch correction
               of gene expression and tissue morphology through
               over-parameterization. We apply STACI to analyze the
               spatio-temporal progression of Alzheimer's disease and identify
               the associated nuclear morphometric and coupled gene expression
               features. Collectively, we demonstrate the importance of
               characterizing disease progression by integrating multiple data
               modalities and its potential for the discovery of disease
               biomarkers.",
  journal   = "Nat. Commun.",
  publisher = "Springer Science and Business Media LLC",
  volume    =  13,
  number    =  1,
  pages     = "7480",
  month_fake     =  dec,
  year      =  2022,
  copyright = "https://creativecommons.org/licenses/by/4.0",
  language  = "en"
}


@ARTICLE{Yang2023-wb,
  title     = "Multi-model adaptive fusion-based graph network for Alzheimer's
               disease prediction",
  author    = "Yang, Fusheng and Wang, Huabin and Wei, Shicheng and Sun,
               Guangming and Chen, Yonglin and Tao, Liang",
  abstract  = "Alzheimer's disease (AD) is a common cognitive disorder.
               Recently, many computer-aided diagnostic techniques have been
               used for AD prediction utilizing deep learning technology, among
               which graph neural networks have received increasing attention
               owing to their ability to model sample relationships on large
               population graphs. Most of the existing graph-based methods
               predict diseases according to a single model, which makes it
               difficult to select an appropriate node embedding algorithm for
               a certain classification task. Moreover, integrating data from
               different patterns into a unified model to improve the quality
               of disease diagnosis remains a challenge. Hence, in this study,
               we aimed to develop a multi-model fusion framework for AD
               prediction. A spectral graph attention model was used to
               aggregate intra- and inter-cluster node embeddings of normal and
               diseased populations, whereafter, a bilinear aggregation model
               was applied as an auxiliary model to enhance the abnormality
               degree in different categories of populations, and finally, an
               adaptive fusion module was designed to dynamically fuse the
               results of both models and enhance AD prediction. Compared to
               other comparison methods, the model proposed in this study
               provides the best results.",
  journal   = "Comput. Biol. Med.",
  publisher = "Elsevier BV",
  volume    =  153,
  number    =  106518,
  pages     = "106518",
  month_fake     =  feb,
  year      =  2023,
  keywords  = "Alzheimer's disease prediction; Computational medicine; Disease
               prediction algorithm; Multi-model fusion",
  language  = "en"
}

@ARTICLE{Bi2022-cx,
  title     = "Feature aggregation graph convolutional network based on imaging
               genetic data for diagnosis and pathogeny identification of
               Alzheimer's disease",
  author    = "Bi, Xia-An and Zhou, Wenyan and Luo, Sheng and Mao, Yuhua and
               Hu, Xi and Zeng, Bin and Xu, Luyun",
  abstract  = "The roles of brain regions activities and gene expressions in
               the development of Alzheimer's disease (AD) remain unclear.
               Existing imaging genetic studies usually has the problem of
               inefficiency and inadequate fusion of data. This study proposes
               a novel deep learning method to efficiently capture the
               development pattern of AD. First, we model the interaction
               between brain regions and genes as node-to-node feature
               aggregation in a brain region-gene network. Second, we propose a
               feature aggregation graph convolutional network (FAGCN) to
               transmit and update the node feature. Compared with the trivial
               graph convolutional procedure, we replace the input from the
               adjacency matrix with a weight matrix based on correlation
               analysis and consider common neighbor similarity to discover
               broader associations of nodes. Finally, we use a full-gradient
               saliency graph mechanism to score and extract the pathogenetic
               brain regions and risk genes. According to the results, FAGCN
               achieved the best performance among both traditional and
               cutting-edge methods and extracted AD-related brain regions and
               genes, providing theoretical and methodological support for the
               research of related diseases.",
  journal   = "Brief. Bioinform.",
  publisher = "Oxford University Press (OUP)",
  volume    =  23,
  number    =  3,
  month_fake     =  may,
  year      =  2022,
  keywords  = "Alzheimer's disease; deep learning; feature aggregation graph
               convolutional network; imaging genetics",
  copyright = "https://academic.oup.com/journals/pages/open\_access/funder\_policies/chorus/standard\_publication\_model",
  language  = "en"
}


%%%%%%%%%%%%%%%%%% multimodal  learning 
@ARTICLE{Baltrusaitis2019-kx,
  title     = "Multimodal machine learning: A survey and taxonomy",
  author    = "Baltrusaitis, Tadas and Ahuja, Chaitanya and Morency,
               Louis-Philippe",
  abstract  = "Our experience of the world is multimodal - we see objects, hear
               sounds, feel texture, smell odors, and taste flavors. Modality
               refers to the way in which something happens or is experienced
               and a research problem is characterized as multimodal when it
               includes multiple such modalities. In order for Artificial
               Intelligence to make progress in understanding the world around
               us, it needs to be able to interpret such multimodal signals
               together. Multimodal machine learning aims to build models that
               can process and relate information from multiple modalities. It
               is a vibrant multi-disciplinary field of increasing importance
               and with extraordinary potential. Instead of focusing on
               specific multimodal applications, this paper surveys the recent
               advances in multimodal machine learning itself and presents them
               in a common taxonomy. We go beyond the typical early and late
               fusion categorization and identify broader challenges that are
               faced by multimodal machine learning, namely: representation,
               translation, alignment, fusion, and co-learning. This new
               taxonomy will enable researchers to better understand the state
               of the field and identify directions for future research.",
  journal   = "IEEE Trans. Pattern Anal. Mach. Intell.",
  publisher = "Institute of Electrical and Electronics Engineers (IEEE)",
  volume    =  41,
  number    =  2,
  pages     = "423--443",
  month_fake     =  feb,
  year      =  2019,
  copyright = "https://ieeexplore.ieee.org/Xplorehelp/downloads/license-information/IEEE.html",
  language  = "en"
}

@ARTICLE{Teipel2015-uz,
  title     = "Multimodal imaging in Alzheimer's disease: validity and
               usefulness for early detection",
  author    = "Teipel, Stefan and Drzezga, Alexander and Grothe, Michel J and
               Barthel, Henryk and Ch{\'e}telat, Ga{\"e}l and Schuff, Norbert
               and Skudlarski, Pawel and Cavedo, Enrica and Frisoni, Giovanni B
               and Hoffmann, Wolfgang and Thyrian, Jochen Ren{\'e} and Fox,
               Chris and Minoshima, Satoshi and Sabri, Osama and Fellgiebel,
               Andreas",
  abstract  = "Alzheimer's disease is a progressive neurodegenerative disease
               that typically manifests clinically as an isolated amnestic
               deficit that progresses to a characteristic dementia syndrome.
               Advances in neuroimaging research have enabled mapping of
               diverse molecular, functional, and structural aspects of
               Alzheimer's disease pathology in ever increasing temporal and
               regional detail. Accumulating evidence suggests that distinct
               types of imaging abnormalities related to Alzheimer's disease
               follow a consistent trajectory during pathogenesis of the
               disease, and that the first changes can be detected years before
               the disease manifests clinically. These findings have fuelled
               clinical interest in the use of specific imaging markers for
               Alzheimer's disease to predict future development of dementia in
               patients who are at risk. The potential clinical usefulness of
               single or multimodal imaging markers is being investigated in
               selected patient samples from clinical expert centres, but
               additional research is needed before these promising imaging
               markers can be successfully translated from research into
               clinical practice in routine care.",
  journal   = "Lancet Neurol.",
  publisher = "Elsevier BV",
  volume    =  14,
  number    =  10,
  pages     = "1037--1053",
  month_fake     =  oct,
  year      =  2015,
  language  = "en"
}

@ARTICLE{Lee2022-zj,
  title     = "Multimodal brain and retinal imaging of dopaminergic
               degeneration in Parkinson disease",
  author    = "Lee, Jee-Young and Martin-Bastida, Antonio and Murueta-Goyena,
               Ane and Gabilondo, I{\~n}igo and Cuenca, Nicol{\'a}s and
               Piccini, Paola and Jeon, Beomseok",
  abstract  = "Parkinson disease (PD) is a progressive disorder characterized
               by dopaminergic neurodegeneration in the brain. The development
               of parkinsonism is preceded by a long prodromal phase, and >50\%
               of dopaminergic neurons can be lost from the substantia nigra by
               the time of the initial diagnosis. Therefore, validation of in
               vivo imaging biomarkers for early diagnosis and monitoring of
               disease progression is essential for future therapeutic
               developments. PET and single-photon emission CT targeting the
               presynaptic terminals of dopaminergic neurons can be used for
               early diagnosis by detecting axonal degeneration in the
               striatum. However, these techniques poorly differentiate
               atypical parkinsonian syndromes from PD, and their availability
               is limited in clinical settings. Advanced MRI in which
               pathological changes in the substantia nigra are visualized with
               diffusion, iron-sensitive susceptibility and
               neuromelanin-sensitive sequences potentially represents a more
               accessible imaging tool. Although these techniques can visualize
               the classic degenerative changes in PD, they might be
               insufficient for phenotyping or prognostication of heterogeneous
               aspects of PD resulting from extranigral pathologies. The retina
               is an emerging imaging target owing to its pathological
               involvement early in PD, which correlates with brain pathology.
               Retinal optical coherence tomography (OCT) is a non-invasive
               technique to visualize structural changes in the retina.
               Progressive parafoveal thinning and fovea avascular zone
               remodelling, as revealed by OCT, provide potential biomarkers
               for early diagnosis and prognostication in PD. As we discuss in
               this Review, multimodal imaging of the substantia nigra and
               retina is a promising tool to aid diagnosis and management of
               PD.",
  journal   = "Nat. Rev. Neurol.",
  publisher = "Springer Science and Business Media LLC",
  volume    =  18,
  number    =  4,
  pages     = "203--220",
  month_fake     =  apr,
  year      =  2022,
  language  = "en"
}

@ARTICLE{Qiu2022-gz,
  title     = "Multimodal deep learning for Alzheimer's disease dementia
               assessment",
  author    = "Qiu, Shangran and Miller, Matthew I and Joshi, Prajakta S and
               Lee, Joyce C and Xue, Chonghua and Ni, Yunruo and Wang, Yuwei
               and De Anda-Duran, Ileana and Hwang, Phillip H and Cramer,
               Justin A and Dwyer, Brigid C and Hao, Honglin and Kaku, Michelle
               C and Kedar, Sachin and Lee, Peter H and Mian, Asim Z and
               Murman, Daniel L and O'Shea, Sarah and Paul, Aaron B and
               Saint-Hilaire, Marie-Helene and Alton Sartor, E and Saxena,
               Aneeta R and Shih, Ludy C and Small, Juan E and Smith,
               Maximilian J and Swaminathan, Arun and Takahashi, Courtney E and
               Taraschenko, Olga and You, Hui and Yuan, Jing and Zhou, Yan and
               Zhu, Shuhan and Alosco, Michael L and Mez, Jesse and Stein, Thor
               D and Poston, Kathleen L and Au, Rhoda and Kolachalama, Vijaya B",
  abstract  = "Worldwide, there are nearly 10 million new cases of dementia
               annually, of which Alzheimer's disease (AD) is the most common.
               New measures are needed to improve the diagnosis of individuals
               with cognitive impairment due to various etiologies. Here, we
               report a deep learning framework that accomplishes multiple
               diagnostic steps in successive fashion to identify persons with
               normal cognition (NC), mild cognitive impairment (MCI), AD, and
               non-AD dementias (nADD). We demonstrate a range of models
               capable of accepting flexible combinations of routinely
               collected clinical information, including demographics, medical
               history, neuropsychological testing, neuroimaging, and
               functional assessments. We then show that these frameworks
               compare favorably with the diagnostic accuracy of practicing
               neurologists and neuroradiologists. Lastly, we apply
               interpretability methods in computer vision to show that
               disease-specific patterns detected by our models track distinct
               patterns of degenerative changes throughout the brain and
               correspond closely with the presence of neuropathological
               lesions on autopsy. Our work demonstrates methodologies for
               validating computational predictions with established standards
               of medical diagnosis.",
  journal   = "Nat. Commun.",
  publisher = "Springer Science and Business Media LLC",
  volume    =  13,
  number    =  1,
  pages     = "3404",
  month_fake     =  jun,
  year      =  2022,
  copyright = "https://creativecommons.org/licenses/by/4.0",
  language  = "en"
}

@ARTICLE{Ruppert2020-zv,
  title     = "Network degeneration in Parkinson's disease: multimodal imaging
               of nigro-striato-cortical dysfunction",
  author    = "Ruppert, Marina C and Greuel, Andrea and Tahmasian, Masoud and
               Schwartz, Frank and St{\"u}rmer, Sophie and Maier, Franziska and
               Hammes, Jochen and Tittgemeyer, Marc and Timmermann, Lars and
               van Eimeren, Thilo and Drzezga, Alexander and Eggers, Carsten",
  abstract  = "The spreading hypothesis of neurodegeneration assumes an
               expansion of neural pathologies along existing neural pathways.
               Multimodal neuroimaging studies have demonstrated distinct
               topographic patterns of cerebral pathologies in
               neurodegeneration. For Parkinson's disease the hypothesis so far
               rests largely on histopathological evidence of
               $\alpha$-synuclein spreading in a characteristic pattern and
               progressive nigrostriatal dopamine depletion. Functional
               consequences of nigrostriatal dysfunction on cortical activity
               remain to be elucidated. Our goal was to investigate multimodal
               imaging correlates of degenerative processes in Parkinson's
               disease by assessing dopamine depletion and its potential effect
               on striatocortical connectivity networks and cortical metabolism
               in relation to parkinsonian symptoms. We combined 18F-DOPA-PET,
               18F-fluorodeoxyglucose (FDG)-PET and resting state functional
               MRI to multimodally characterize network alterations in
               Parkinson's disease. Forty-two patients with mild-to-moderate
               stage Parkinson's disease and 14 age-matched healthy control
               subjects underwent a multimodal imaging protocol and
               comprehensive clinical examination. A voxel-wise group
               comparison of 18F-DOPA uptake identified the exact location and
               extent of putaminal dopamine depletion in patients. Resulting
               clusters were defined as seeds for a seed-to-voxel functional
               connectivity analysis. 18F-FDG metabolism was compared between
               groups at a whole-brain level and uptake values were extracted
               from regions with reduced putaminal connectivity. To unravel
               associations between dopaminergic activity, striatocortical
               connectivity, glucose metabolism and symptom severity,
               correlations between normalized uptake values, seed-to-cluster
               $\beta$-values and clinical parameters were tested while
               controlling for age and dopaminergic medication. Aside from
               cortical hypometabolism, 18F-FDG-PET data for the first time
               revealed a hypometabolic midbrain cluster in patients with
               Parkinson's disease that comprised caudal parts of the bilateral
               substantia nigra pars compacta. Putaminal dopamine synthesis
               capacity was significantly reduced in the bilateral posterior
               putamen and correlated with ipsilateral nigral 18F-FDG uptake.
               Resting state functional MRI data indicated significantly
               reduced functional connectivity between the dopamine depleted
               putaminal seed and cortical areas primarily belonging to the
               sensorimotor network in patients with Parkinson's disease. In
               the inferior parietal cortex, hypoconnectivity in patients was
               significantly correlated with lower metabolism (left P = 0.021,
               right P = 0.018). Of note, unilateral network alterations
               quantified with different modalities corresponded with
               contralateral motor impairments. In conclusion, our results
               support the hypothesis that degeneration of nigrostriatal fibres
               functionally impairs distinct striatocortical connections,
               disturbing the efficient interplay between motor processing
               areas and impairing motor control in patients with Parkinson's
               disease. The present study is the first to reveal trimodal
               evidence for network-dependent degeneration in Parkinson's
               disease by outlining the impact of functional nigrostriatal
               pathway impairment on striatocortical functional connectivity
               networks and cortical metabolism.",
  journal   = "Brain",
  publisher = "Oxford University Press (OUP)",
  volume    =  143,
  number    =  3,
  pages     = "944--959",
  month_fake     =  mar,
  year      =  2020,
  keywords  = "Parkinson's disease; multimodal imaging; network degeneration;
               neurodegeneration",
  copyright = "https://academic.oup.com/journals/pages/open\_access/funder\_policies/chorus/standard\_publication\_model",
  language  = "en"
}

@ARTICLE{Azam2022-tu,
  title     = "A review on multimodal medical image fusion: Compendious
               analysis of medical modalities, multimodal databases, fusion
               techniques and quality metrics",
  author    = "Azam, Muhammad Adeel and Khan, Khan Bahadar and Salahuddin, Sana
               and Rehman, Eid and Khan, Sajid Ali and Khan, Muhammad Attique
               and Kadry, Seifedine and Gandomi, Amir H",
  abstract  = "BACKGROUND AND OBJECTIVES: Over the past two decades, medical
               imaging has been extensively apply to diagnose diseases. Medical
               experts continue to have difficulties for diagnosing diseases
               with a single modality owing to a lack of information in this
               domain. Image fusion may be use to merge images of specific
               organs with diseases from a variety of medical imaging systems.
               Anatomical and physiological data may be included in
               multi-modality image fusion, making diagnosis simpler. It is a
               difficult challenge to find the best multimodal medical database
               with fusion quality evaluation for assessing recommended image
               fusion methods. As a result, this article provides a complete
               overview of multimodal medical image fusion methodologies,
               databases, and quality measurements. METHODS: In this article, a
               compendious review of different medical imaging modalities and
               evaluation of related multimodal databases along with the
               statistical results is provided. The medical imaging modalities
               are organized based on radiation, visible-light imaging,
               microscopy, and multimodal imaging. RESULTS: The medical imaging
               acquisition is categorized into invasive or non-invasive
               techniques. The fusion techniques are classified into six main
               categories: frequency fusion, spatial fusion, decision-level
               fusion, deep learning, hybrid fusion, and sparse representation
               fusion. In addition, the associated diseases for each modality
               and fusion approach presented. The quality assessments fusion
               metrics are also encapsulated in this article. CONCLUSIONS: This
               survey provides a baseline guideline to medical experts in this
               technical domain that may combine preoperative, intraoperative,
               and postoperative imaging, Multi-sensor fusion for disease
               detection, etc. The advantages and drawbacks of the current
               literature are discussed, and future insights are provided
               accordingly.",
  journal   = "Comput. Biol. Med.",
  publisher = "Elsevier BV",
  volume    =  144,
  number    =  105253,
  pages     = "105253",
  month_fake     =  may,
  year      =  2022,
  keywords  = "Fusion techniques; Image fusion quality metrics; Multimodal
               databases; Multimodal medical image fusion",
  language  = "en"
}

@ARTICLE{Cui2023-kw,
  title    = "Deep multimodal fusion of image and non-image data in disease
              diagnosis and prognosis: a review",
  author   = "Cui, Can and Yang, Haichun and Wang, Yaohong and Zhao, Shilin and
              Asad, Zuhayr and Coburn, Lori A and Wilson, Keith T and Landman,
              Bennett A and Huo, Yuankai",
  abstract = "The rapid development of diagnostic technologies in healthcare is
              leading to higher requirements for physicians to handle and
              integrate the heterogeneous, yet complementary data that are
              produced during routine practice. For instance, the personalized
              diagnosis and treatment planning for a single cancer patient
              relies on various images (e.g. radiology, pathology and camera
              images) and non-image data (e.g. clinical data and genomic data).
              However, such decision-making procedures can be subjective,
              qualitative, and have large inter-subject variabilities. With the
              recent advances in multimodal deep learning technologies, an
              increasingly large number of efforts have been devoted to a key
              question: how do we extract and aggregate multimodal information
              to ultimately provide more objective, quantitative computer-aided
              clinical decision making? This paper reviews the recent studies
              on dealing with such a question. Briefly, this review will
              include the (a) overview of current multimodal learning
              workflows, (b) summarization of multimodal fusion methods, (c)
              discussion of the performance, (d) applications in disease
              diagnosis and prognosis, and (e) challenges and future
              directions.",
  journal  = "Prog. Biomed. Eng. (Bristol)",
  volume   =  5,
  number   =  2,
  month_fake    =  apr,
  year     =  2023,
  keywords = "diagnosis and prognosis; fusion method; medical data; multimodal
              learning",
  language = "en"
}

@ARTICLE{Pawlowski2023-nl,
  title     = "Effective techniques for multimodal data fusion: A comparative
               analysis",
  author    = "Paw{\l}owski, Maciej and Wr{\'o}blewska, Anna and
               Sysko-Roma{\'n}czuk, Sylwia",
  abstract  = "Data processing in robotics is currently challenged by the
               effective building of multimodal and common representations.
               Tremendous volumes of raw data are available and their smart
               management is the core concept of multimodal learning in a new
               paradigm for data fusion. Although several techniques for
               building multimodal representations have been proven successful,
               they have not yet been analyzed and compared in a given
               production setting. This paper explored three of the most common
               techniques, (1) the late fusion, (2) the early fusion, and (3)
               the sketch, and compared them in classification tasks. Our paper
               explored different types of data (modalities) that could be
               gathered by sensors serving a wide range of sensor applications.
               Our experiments were conducted on Amazon Reviews, MovieLens25M,
               and Movie-Lens1M datasets. Their outcomes allowed us to confirm
               that the choice of fusion technique for building multimodal
               representation is crucial to obtain the highest possible model
               performance resulting from the proper modality combination.
               Consequently, we designed criteria for choosing this optimal
               data fusion technique.",
  journal   = "Sensors (Basel)",
  publisher = "MDPI AG",
  volume    =  23,
  number    =  5,
  pages     = "2381",
  month_fake     =  feb,
  year      =  2023,
  keywords  = "comparative analysis; data fusion; deep learning in sensor
               systems; multimodal learning; multimodal representation; neural
               networks",
  copyright = "https://creativecommons.org/licenses/by/4.0/",
  language  = "en"
}

@ARTICLE{Pereira2023-dc,
  title     = "A comparative analysis of early and late fusion for the
               multimodal two-class problem",
  author    = "Pereira, Luis Manuel and Salazar, Addisson and Vergara, Luis",
  journal   = "IEEE Access",
  publisher = "Institute of Electrical and Electronics Engineers (IEEE)",
  volume    =  11,
  pages     = "84283--84300",
  year      =  2023,
  copyright = "https://creativecommons.org/licenses/by-nc-nd/4.0/"
}
%%%%%%%%%%%% hyperbolic

@inproceedings{pmlr-v202-mishne23a,
  title = 	 "The Numerical Stability of Hyperbolic Representation Learning",
  author =       "Mishne, Gal and Wan, Zhengchao and Wang, Yusu and Yang, Sheng",
  booktitle = 	 "Proceedings of the 40th International Conference on Machine Learning",
  pages = 	 "24925--24949",
  year = 	 2023,
  editor = 	 "Krause, Andreas and Brunskill, Emma and Cho, Kyunghyun and Engelhardt, Barbara and Sabato, Sivan and Scarlett, Jonathan",
  volume = 	 202,
  series = 	 "Proceedings of Machine Learning Research",
  month_fake = 	 "23--29 Jul",
  publisher =    "PMLR",
  pdf = 	 "https://proceedings.mlr.press/v202/mishne23a/mishne23a.pdf",
  url = 	 "https://proceedings.mlr.press/v202/mishne23a.html",
  abstract = 	 "The hyperbolic space is widely used for representing hierarchical datasets due to its ability to embed trees with small distortion. However, this property comes at a price of numerical instability such that training hyperbolic learning models will sometimes lead to catastrophic NaN problems, encountering unrepresentable values in floating point arithmetic. In this work, we analyze the limitations of two popular models for the hyperbolic space, namely, the Poincaré ball and the Lorentz model. We find that, under the 64-bit arithmetic system, the Poincaré ball has a relatively larger capacity than the Lorentz model for correctly representing points. However, the Lorentz model is superior to the Poincaré ball from the perspective of optimization, which we theoretically validate. To address these limitations, we identify one Euclidean parametrization of the hyperbolic space which can alleviate these issues. We further extend this Euclidean parametrization to hyperbolic hyperplanes and demonstrate its effectiveness in improving the performance of hyperbolic SVM."
}

@ARTICLE{Ganea2018-ct,
  title="Hyperbolic neural networks",
  author="Ganea, Octavian and B{\'e}cigneul, Gary and Hofmann, Thomas",
  journal="Advances in neural information processing systems",
  volume=31,
  year=2018
}


%%%%%%%%%%%%%%%%%% hyperbolic for brain 
@ARTICLE{Baker2023-zg,
  title    = "Hyperbolic graph embedding of {MEG} brain networks to study brain
              alterations in individuals with subjective cognitive decline",
  author   = "Baker, Cole and Su{\'a}rez-M{\'e}ndez, Isabel and Smith, Grace
              and Marsh, Elisabeth B and Funke, Michael and Mosher, John C and
              Maest{\'u}, Fernando and Xu, Mengjia and Pantazis, Dimitrios",
  abstract = "AbstractAn expansive area of research focuses on discerning
              patterns of alterations in functional brain networks from the
              early stages of Alzheimer's disease, even at the subjective
              cognitive decline (SCD) stage. Here, we developed a novel
              hyperbolic MEG brain network embedding framework for transforming
              high-dimensional complex MEG brain networks into
              lower-dimensional hyperbolic representations. Using this model,
              we computed hyperbolic embeddings of the MEG brain networks of
              two distinct participant groups: individuals with SCD and healthy
              controls. We demonstrated that these embeddings preserve both
              local and global geometric information, presenting reduced
              distortion compared to rival models, even when brain networks are
              mapped into low-dimensional spaces. In addition, our findings
              showed that the hyperbolic embeddings encompass unique
              SCD-related information that improves the discriminatory power
              above and beyond that of connectivity features alone. Notably, we
              introduced a unique metric---the radius of the node
              embeddings---which effectively proxies the hierarchical
              organization of the brain. Using this metric, we identified
              subtle hierarchy organizational differences between the two
              participant groups, suggesting increased hierarchy in the dorsal
              attention, frontoparietal, and ventral attention subnetworks
              among the SCD group. Last, we assessed the correlation between
              these hierarchical variations and cognitive assessment scores,
              revealing associations with diminished performance across
              multiple cognitive evaluations in the SCD group. Overall, this
              study presents the first evaluation of hyperbolic embeddings of
              MEG brain networks, offering novel insights into brain
              organization, cognitive decline, and potential diagnostic avenues
              of Alzheimer's disease.",
  journal  = "bioRxiv",
  month_fake    =  oct,
  year     =  2023,
  publisher="Cold Spring Harbor Laboratory Preprints"
}


@ARTICLE{Whi2022-za,
  title     = "Hyperbolic disc embedding of functional human brain connectomes
               using resting-state {fMRI}",
  author    = "Whi, Wonseok and Ha, Seunggyun and Kang, Hyejin and Lee, Dong
               Soo",
  abstract  = "The brain presents a real complex network of modular,
               small-world, and hierarchical nature, which are features of
               non-Euclidean geometry. Using resting-state functional magnetic
               resonance imaging, we constructed a scale-free binary graph for
               each subject, using internodal time series correlation of
               regions of interest as a proximity measure. The resulting
               network could be embedded onto manifolds of various curvatures
               and dimensions. While maintaining the fidelity of embedding (low
               distortion, high mean average precision), functional brain
               networks were found to be best represented in the hyperbolic
               disc. Using the 𝕊1/ℍ2 model, we reduced the dimension of the
               network into two-dimensional hyperbolic space and were able to
               efficiently visualize the internodal connections of the brain,
               preserving proximity as distances and angles on the hyperbolic
               discs. Each individual disc revealed relevance with its anatomic
               counterpart and absence of center-spaced node. Using the
               hyperbolic distance on the 𝕊1/ℍ2 model, we could detect the
               anomaly of network in autism spectrum disorder subjects. This
               procedure of embedding grants us a reliable new framework for
               studying functional brain networks and the possibility of
               detecting anomalies of the network in the hyperbolic disc on an
               individual scale.",
  journal   = "Netw. Neurosci.",
  publisher = "MIT Press - Journals",
  volume    =  6,
  number    =  3,
  pages     = "745--764",
  month_fake     =  jul,
  year      =  2022,
  keywords  = "Functional brain network; Hyperbolic embedding; Hyperbolic
               geometry; 𝕊1/ℍ2 model",
  copyright = "https://creativecommons.org/licenses/by/4.0/",
  language  = "en"
}

@INCOLLECTION{Zhang2023-lw,
  title     = "Multimodal deep fusion in hyperbolic space for mild cognitive
               impairment study",
  booktitle = "Lecture Notes in Computer Science",
  author    = "Zhang, Lu and Na, Saiyang and Liu, Tianming and Zhu, Dajiang and
               Huang, Junzhou",
  publisher = "Springer Nature Switzerland",
  pages     = "674--684",
  series    = "Lecture notes in computer science",
  year      =  2023,
  address   = "Cham",
  copyright = "https://www.springernature.com/gp/researchers/text-and-data-mining"
}


@ARTICLE{Zheng2020-tw,
  title     = "Geometric renormalization unravels self-similarity of the
               multiscale human connectome",
  author    = "Zheng, Muhua and Allard, Antoine and Hagmann, Patric and
               Alem{\'a}n-G{\'o}mez, Yasser and Serrano, M {\'A}ngeles",
  abstract  = "Structural connectivity in the brain is typically studied by
               reducing its observation to a single spatial resolution.
               However, the brain possesses a rich architecture organized over
               multiple scales linked to one another. We explored the
               multiscale organization of human connectomes using datasets of
               healthy subjects reconstructed at five different resolutions. We
               found that the structure of the human brain remains self-similar
               when the resolution of observation is progressively decreased by
               hierarchical coarse-graining of the anatomical regions.
               Strikingly, a geometric network model, where distances are not
               Euclidean, predicts the multiscale properties of connectomes,
               including self-similarity. The model relies on the application
               of a geometric renormalization protocol which decreases the
               resolution by coarse-graining and averaging over short
               similarity distances. Our results suggest that simple organizing
               principles underlie the multiscale architecture of human
               structural brain networks, where the same connectivity law
               dictates short- and long-range connections between different
               brain regions over many resolutions. The implications are varied
               and can be substantial for fundamental debates, such as whether
               the brain is working near a critical point, as well as for
               applications including advanced tools to simplify the digital
               reconstruction and simulation of the brain.",
  journal   = "Proc. Natl. Acad. Sci. U. S. A.",
  publisher = "Proceedings of the National Academy of Sciences",
  volume    =  117,
  number    =  33,
  pages     = "20244--20253",
  month_fake     =  aug,
  year      =  2020,
  keywords  = "human brain; multiscale structure; network geometry;
               neuroscience; renormalization",
  copyright = "https://www.pnas.org/site/aboutpnas/licenses.xhtml",
  language  = "en"
}

@ARTICLE{Sharpee2019-nb,
  title     = "An argument for hyperbolic geometry in neural circuits",
  author    = "Sharpee, Tatyana O",
  journal   = "Curr. Opin. Neurobiol.",
  publisher = "Elsevier BV",
  volume    =  58,
  pages     = "101--104",
  month_fake     =  oct,
  year      =  2019,
  language  = "en"
}

@ARTICLE{Zhang2023-vq,
  title    = "Hippocampal spatial representations exhibit a hyperbolic geometry
              that expands with experience",
  author   = "Zhang, Huanqiu and Rich, P Dylan and Lee, Albert K and Sharpee,
              Tatyana O",
  abstract = "Daily experience suggests that we perceive distances near us
              linearly. However, the actual geometry of spatial representation
              in the brain is unknown. Here we report that neurons in the CA1
              region of rat hippocampus that mediate spatial perception
              represent space according to a non-linear hyperbolic geometry.
              This geometry uses an exponential scale and yields greater
              positional information than a linear scale. We found that the
              size of the representation matches the optimal predictions for
              the number of CA1 neurons. The representations also dynamically
              expanded proportional to the logarithm of time that the animal
              spent exploring the environment, in correspondence with the
              maximal mutual information that can be received. The dynamic
              changes tracked even small variations due to changes in the
              running speed of the animal. These results demonstrate how neural
              circuits achieve efficient representations using dynamic
              hyperbolic geometry.",
  journal  = "Nat. Neurosci.",
  volume   =  26,
  number   =  1,
  pages    = "131--139",
  month_fake    =  jan,
  year     =  2023,
  language = "en"
}

@ARTICLE{Tadic2019-wo,
  title     = "Functional geometry of human connectomes",
  author    = "Tadi{\'c}, Bosiljka and Andjelkovi{\'c}, Miroslav and Melnik,
               Roderick",
  abstract  = "Mapping the brain imaging data to networks, where nodes
               represent anatomical brain regions and edges indicate the
               occurrence of fiber tracts between them, has enabled an
               objective graph-theoretic analysis of human connectomes.
               However, the latent structure on higher-order interactions
               remains unexplored, where many brain regions act in synergy to
               perform complex functions. Here we use the simplicial complexes
               description of human connectome, where the shared simplexes
               encode higher-order relationships between groups of nodes. We
               study consensus connectome of 100 female (F-connectome) and of
               100 male (M-connectome) subjects that we generated from the
               Budapest Reference Connectome Server v3.0 based on data from the
               Human Connectome Project. Our analysis reveals that the
               functional geometry of the common F\&M-connectome coincides with
               the M-connectome and is characterized by a complex architecture
               of simplexes to the 14th order, which is built in six anatomical
               communities, and linked by short cycles. The F-connectome has
               additional edges that involve different brain regions, thereby
               increasing the size of simplexes and introducing new cycles.
               Both connectomes contain characteristic subjacent graphs that
               make them 3/2-hyperbolic. These results shed new light on the
               functional architecture of the brain, suggesting that insightful
               differences among connectomes are hidden in their higher-order
               connectivity.",
  journal   = "Sci. Rep.",
  publisher = "Springer Science and Business Media LLC",
  volume    =  9,
  number    =  1,
  pages     = "12060",
  month_fake     =  aug,
  year      =  2019,
  copyright = "https://creativecommons.org/licenses/by/4.0",
  language  = "en"
}

@ARTICLE{Whi2022-lm,
  title     = "Characteristic functional cores revealed by hyperbolic disc
               embedding and k-core percolation on resting-state {fMRI}",
  author    = "Whi, Wonseok and Huh, Youngmin and Ha, Seunggyun and Lee,
               Hyekyoung and Kang, Hyejin and Lee, Dong Soo",
  abstract  = "Hyperbolic disc embedding and k-core percolation reveal the
               hierarchical structure of functional connectivity on
               resting-state fMRI (rsfMRI). Using 180 normal adults' rsfMRI
               data from the human connectome project database, we visualized
               inter-voxel relations by embedding voxels on the hyperbolic
               space using the [Formula: see text] model. We also conducted
               k-core percolation on 30 participants to investigate core voxels
               for each individual. It recursively peels the layer off, and
               this procedure leaves voxels embedded in the center of the
               hyperbolic disc. We used independent components to classify core
               voxels, and it revealed stereotypes of individuals such as
               visual network dominant, default mode network dominant, and
               distributed patterns. Characteristic core structures of
               resting-state brain connectivity of normal subjects disclosed
               the distributed or asymmetric contribution of voxels to the
               kmax-core, which suggests the hierarchical dominance of certain
               IC subnetworks characteristic of subgroups of individuals at
               rest.",
  journal   = "Sci. Rep.",
  publisher = "Springer Science and Business Media LLC",
  volume    =  12,
  number    =  1,
  pages     = "4887",
  month_fake     =  mar,
  year      =  2022,
  copyright = "https://creativecommons.org/licenses/by/4.0",
  language  = "en"
}


% The entry below contains non-ASCII chars that could not be converted
% to a LaTeX equivalent.


%%%%%%%% GCN 10.48550/arXiv.1609.02907

@inproceedings{Kipf2016-ql,
  title={Semi-Supervised Classification with Graph Convolutional Networks},
  author={Thomas N. Kipf and Max Welling},
  booktitle={International Conference on Learning Representations},
  year={2017},
  url={https://openreview.net/forum?id=SJU4ayYgl}
}


@InProceedings{pmlr-v119-lou20a,
  title = 	 {Differentiating through the Fr{é}chet Mean},
  author =       {Lou, Aaron and Katsman, Isay and Jiang, Qingxuan and Belongie, Serge and Lim, Ser-Nam and De Sa, Christopher},
  booktitle = 	 {Proceedings of the 37th International Conference on Machine Learning},
  pages = 	 {6393--6403},
  year = 	 {2020},
  editor = 	 {III, Hal Daumé and Singh, Aarti},
  volume = 	 {119},
  series = 	 {Proceedings of Machine Learning Research},
  month_fake = 	 {13--18 Jul},
  publisher =    {PMLR},
  abstract = 	 {Recent advances in deep representation learning on Riemannian manifolds extend classical deep learning operations to better capture the geometry of the manifold. One possible extension is the Fr{é}chet mean, the generalization of the Euclidean mean; however, it has been difficult to apply because it lacks a closed form with an easily computable derivative. In this paper, we show how to differentiate through the Fr{é}chet mean for arbitrary Riemannian manifolds. Then, focusing on hyperbolic space, we derive explicit gradient expressions and a fast, accurate, and hyperparameter-free Fr{é}chet mean solver. This fully integrates the Fr{é}chet mean into the hyperbolic neural network pipeline. To demonstrate this integration, we present two case studies. First, we apply our Fr{é}chet mean to the existing Hyperbolic Graph Convolutional Network, replacing its projected aggregation to obtain state-of-the-art results on datasets with high hyperbolicity. Second, to demonstrate the Fr{é}chet mean’s capacity to generalize Euclidean neural network operations, we develop a hyperbolic batch normalization method that gives an improvement parallel to the one observed in the Euclidean setting.}
}

@ARTICLE{Cao2023-hm,
  title     = "Poincar{\'e} Fr{\'e}chet mean",
  author    = "Cao, Xiaofeng",
  journal   = "Pattern Recognit.",
  publisher = "Elsevier BV",
  volume    =  137,
  number    =  109302,
  pages     = "109302",
  month_fake     =  may,
  year      =  2023,
  language  = "en"
}

@InProceedings{pmlr-v119-chen20j,
  title = 	 {A Simple Framework for Contrastive Learning of Visual Representations},
  author =       {Chen, Ting and Kornblith, Simon and Norouzi, Mohammad and Hinton, Geoffrey},
  booktitle = 	 {Proceedings of the 37th International Conference on Machine Learning},
  pages = 	 {1597--1607},
  year = 	 {2020},
  editor = 	 {III, Hal Daumé and Singh, Aarti},
  volume = 	 {119},
  series = 	 {Proceedings of Machine Learning Research},
  month = 	 {13--18 Jul},
  publisher =    {PMLR},
  pdf = 	 {http://proceedings.mlr.press/v119/chen20j/chen20j.pdf},
  url = 	 {https://proceedings.mlr.press/v119/chen20j.html},
  abstract = 	 {This paper presents SimCLR: a simple framework for contrastive learning of visual representations. We simplify recently proposed contrastive self-supervised learning algorithms without requiring specialized architectures or a memory bank. In order to understand what enables the contrastive prediction tasks to learn useful representations, we systematically study the major components of our framework. We show that (1) composition of data augmentations plays a critical role in defining effective predictive tasks, (2) introducing a learnable nonlinear transformation between the representation and the contrastive loss substantially improves the quality of the learned representations, and (3) contrastive learning benefits from larger batch sizes and more training steps compared to supervised learning. By combining these findings, we are able to considerably outperform previous methods for self-supervised and semi-supervised learning on ImageNet. A linear classifier trained on self-supervised representations learned by SimCLR achieves 76.5% top-1 accuracy, which is a 7% relative improvement over previous state-of-the-art, matching the performance of a supervised ResNet-50. When fine-tuned on only 1% of the labels, we achieve 85.8% top-5 accuracy, outperforming AlexNet with 100X fewer labels.}
}

@InProceedings{Chen2020-au,
  title         = "A simple framework for contrastive learning of visual
                   representations",
  author        = "Chen, Ting and Kornblith, Simon and Norouzi, Mohammad and
                   Hinton, Geoffrey",
  abstract      = "This paper presents SimCLR: a simple framework for
                   contrastive learning of visual representations. We simplify
                   recently proposed contrastive self-supervised learning
                   algorithms without requiring specialized architectures or a
                   memory bank. In order to understand what enables the
                   contrastive prediction tasks to learn useful
                   representations, we systematically study the major
                   components of our framework. We show that (1) composition of
                   data augmentations plays a critical role in defining
                   effective predictive tasks, (2) introducing a learnable
                   nonlinear transformation between the representation and the
                   contrastive loss substantially improves the quality of the
                   learned representations, and (3) contrastive learning
                   benefits from larger batch sizes and more training steps
                   compared to supervised learning. By combining these
                   findings, we are able to considerably outperform previous
                   methods for self-supervised and semi-supervised learning on
                   ImageNet. A linear classifier trained on self-supervised
                   representations learned by SimCLR achieves 76.5\% top-1
                   accuracy, which is a 7\% relative improvement over previous
                   state-of-the-art, matching the performance of a supervised
                   ResNet-50. When fine-tuned on only 1\% of the labels, we
                   achieve 85.8\% top-5 accuracy, outperforming AlexNet with
                   100X fewer labels.",
  month_fake         =  feb,
  year          =  2020,
  journal = "arXiv",
  primaryClass  = "cs.LG",
  eprint        = "2002.05709",
  booktitle = 	"Proceedings of the 37th International Conference on Machine Learning",
  pages = 	"1597--1607",
  volume = 	"119",
  publisher =    "PMLR",
}

@ARTICLE{Sun2024-fd,
  title     = "From electroencephalogram data to brain networks:
               Graph-learning-based brain disease diagnosis",
  author    = "Sun, Ke and Peng, Ciyuan and Yu, Shuo and Han, Zhuoyang and Xia,
               Feng",
  journal   = "IEEE Intell. Syst.",
  publisher = "Institute of Electrical and Electronics Engineers (IEEE)",
  volume    =  39,
  number    =  2,
  pages     = "21--29",
  month     =  mar,
  year      =  2024,
  copyright = "https://ieeexplore.ieee.org/Xplorehelp/downloads/license-information/IEEE.html"
}

@inproceedings{peng2024adaptive,
  title={Adaptive Brain Network Augmentation Based on Group-aware Graph Learning},
  author={Peng, Ciyuan and Liu, Mujie and Meng, Chenxuan and Yu, Shuo and Xia, Feng},
  booktitle={ICLR},
  year={2024}
}

@INCOLLECTION{Febrinanto2023-wt,
  title     = "Balanced graph structure information for brain disease detection",
  booktitle = "Knowledge Management and Acquisition for Intelligent Systems",
  author    = "Febrinanto, Falih Gozi and Liu, Mujie and Xia, Feng",
  publisher = "Springer Nature Singapore",
  pages     = "134--143",
  series    = "Lecture notes in computer science",
  year      =  2023,
  address   = "Singapore",
  copyright = "https://www.springernature.com/gp/researchers/text-and-data-mining"
}


@ARTICLE{Tu2024-di,
  title     = "Deep outdatEd fAct detectioN in knowledge graphs",
  author    = "Tu, Huiling and Yu, Shuo and Saikrishna, Vidya and Xia, Feng and
               Verspoor, Karin",
  abstract  = "Knowledge graphs (KGs) have garnered significant attention for
               their vast potential across diverse domains. However, the issue
               of outdated facts poses a challenge to KGs, affecting their
               overall quality as real-world information evolves. Existing
               solutions for outdated fact detection often rely on manual
               recognition. In response, this paper presents DEAN (Deep
               outdatEd fAct detectioN), a novel deep learning-based framework
               designed to identify outdated facts within KGs. DEAN
               distinguishes itself by capturing implicit structural
               information among facts through comprehensive modeling of both
               entities and relations. To effectively uncover latent
               out-of-date information, DEAN employs a contrastive approach
               based on a pre-defined Relations-to-Nodes (R2N) graph, weighted
               by the number of entities. Experimental results demonstrate the
               effectiveness and superiority of DEAN over state-of-the-art
               baseline methods.",
  publisher = "arXiv",
  year      =  2024
}

@ARTICLE{Yue2023-gq,
  title     = "Hyperbolic Contrastive Learning",
  author    = "Yue, Yun and Lin, Fangzhou and Yamada, Kazunori D and Zhang,
               Ziming",
  abstract  = "Learning good image representations that are beneficial to
               downstream tasks is a challenging task in computer vision. As
               such, a wide variety of self-supervised learning approaches have
               been proposed. Among them, contrastive learning has shown
               competitive performance on several benchmark datasets. The
               embeddings of contrastive learning are arranged on a hypersphere
               that results in using the inner (dot) product as a distance
               measurement in Euclidean space. However, the underlying
               structure of many scientific fields like social networks, brain
               imaging, and computer graphics data exhibit highly non-Euclidean
               latent geometry. We propose a novel contrastive learning
               framework to learn semantic relationships in the hyperbolic
               space. Hyperbolic space is a continuous version of trees that
               naturally owns the ability to model hierarchical structures and
               is thus beneficial for efficient contrastive representation
               learning. We also extend the proposed Hyperbolic Contrastive
               Learning (HCL) to the supervised domain and studied the
               adversarial robustness of HCL. The comprehensive experiments
               show that our proposed method achieves better results on
               self-supervised pretraining, supervised classification, and
               higher robust accuracy than baseline methods.",
  journal = "arXiv preprint arXiv:2302.01409",
  year      =  2023
}

@INPROCEEDINGS{Wang2018-mo,
  title           = "A novel multimodal {MRI} analysis for Alzheimer's disease
                     based on convolutional neural network",
  booktitle       = "2018 40th Annual International Conference of the {IEEE}
                     Engineering in Medicine and Biology Society ({EMBC})",
  author          = "Wang, Yan and Yang, Yanwu and Guo, Xin and Ye, Chenfei and
                     Gao, Na and Fang, Yuan and Ma, Heather T",
  publisher       = "IEEE",
  month_fake           =  jul,
  year            =  2018,
  conference      = "2018 40th Annual International Conference of the IEEE
                     Engineering in Medicine and Biology Society (EMBC)",
  location        = "Honolulu, HI"
}

@ARTICLE{Song2021-ev,
  title     = "Graph convolution network with similarity awareness and adaptive
               calibration for disease-induced deterioration prediction",
  author    = "Song, Xuegang and Zhou, Feng and Frangi, Alejandro F and Cao,
               Jiuwen and Xiao, Xiaohua and Lei, Yi and Wang, Tianfu and Lei,
               Baiying",
  abstract  = "Graph convolution networks (GCN) have been successfully applied
               in disease prediction tasks as they capture interactions (i.e.,
               edges and edge weights on the graph) between individual
               elements. The interactions in existing works are constructed by
               fusing similarity between imaging information and distance
               between non-imaging information, whereas disregarding the
               disease status of those individuals in the training set.
               Besides, the similarity is being evaluated by computing the
               correlation distance between feature vectors, which limits
               prediction performance, especially for predicting significant
               memory concern (SMC) and mild cognitive impairment (MCI). In
               this paper, we propose three mechanisms to improve GCN, namely
               similarity-aware adaptive calibrated GCN (SAC-GCN), for
               predicting SMC and MCI. First, we design a similarity-aware
               graph using different receptive fields to consider disease
               status. The labelled subjects on the graph are only connected
               with those labelled subjects with the same status. Second, we
               propose an adaptive mechanism to evaluate similarity.
               Specifically, we construct initial GCN with evaluating
               similarity by using traditional correlation distance, then
               pre-train the initial GCN by using training samples and use it
               to score all subjects. Then, the difference between these scores
               replaces correlation distance to update similarity. Last, we
               devise a calibration mechanism to fuse functional magnetic
               resonance imaging (fMRI) and diffusion tensor imaging (DTI)
               information into edges. The proposed method is tested on the
               Alzheimer's Disease Neuroimaging Initiative (ADNI) dataset.
               Experimental results demonstrate that our proposed method is
               useful to predict disease-induced deterioration and superior to
               other related algorithms, with a mean classification accuracy of
               86.83\% in our prediction tasks.",
  journal   = "Med. Image Anal.",
  publisher = "Elsevier BV",
  volume    =  69,
  number    =  101947,
  pages     = "101947",
  month_fake     =  apr,
  year      =  2021,
  keywords  = "Adaptive mechanism; Calibration mechanism; Disease prediction;
               Dual-modal information; Graph convolution network; Similarity
               awareness",
  language  = "en"
}

@ARTICLE{Ismail2023-rs,
  title     = "A meta-heuristic multi-objective optimization method for
               Alzheimer's disease detection based on multi-modal data",
  author    = "Ismail, Walaa N and Rajeena, Fathimathul and Ali, Mona A S",
  abstract  = "Alzheimer's disease (AD) is a neurodegenerative disease that
               affects a large number of people across the globe. Even though
               AD is one of the most commonly seen brain disorders, it is
               difficult to detect and it requires a categorical representation
               of features to differentiate similar patterns. Research into
               more complex problems, such as AD detection, frequently employs
               neural networks. Those approaches are regarded as
               well-understood and even sufficient by researchers and
               scientists without formal training in artificial intelligence.
               Thus, it is imperative to identify a method of detection that is
               fully automated and user-friendly to non-AI experts. The method
               should find efficient values for models' design parameters
               promptly to simplify the neural network design process and
               subsequently democratize artificial intelligence. Further,
               multi-modal medical image fusion has richer modal features and a
               superior ability to represent information. A fusion image is
               formed by integrating relevant and complementary information
               from multiple input images to facilitate more accurate diagnosis
               and better treatment. This study presents a MultiAz-Net as a
               novel optimized ensemble-based deep neural network learning
               model that incorporate heterogeneous information from PET and
               MRI images to diagnose Alzheimer's disease. Based on features
               extracted from the fused data, we propose an automated procedure
               for predicting the onset of AD at an early stage. Three steps
               are involved in the proposed architecture: image fusion, feature
               extraction, and classification. Additionally, the
               Multi-Objective Grasshopper Optimization Algorithm (MOGOA) is
               presented as a multi-objective optimization algorithm to
               optimize the layers of the MultiAz-Net. The desired objective
               functions are imposed to achieve this, and the design parameters
               are searched for corresponding values. The proposed deep
               ensemble model has been tested to perform four Alzheimer's
               disease categorization tasks, three binary categorizations, and
               one multi-class categorization task by utilizing the publicly
               available Alzheimer neuroimaging dataset. The proposed method
               achieved (92.3 $\pm$ 5.45)\% accuracy for the
               multi-class-classification task, significantly better than other
               network models that have been reported.",
  journal   = "Mathematics",
  publisher = "MDPI AG",
  volume    =  11,
  number    =  4,
  pages     = "957",
  month_fake     =  feb,
  year      =  2023,
  copyright = "https://creativecommons.org/licenses/by/4.0/",
  language  = "en"
}

@ARTICLE{Kong2022-lj,
  title     = "Multi-modal data Alzheimer's disease detection based on {3D}
               convolution",
  author    = "Kong, Zhaokai and Zhang, Mengyi and Zhu, Wenjun and Yi, Yang and
               Wang, Tian and Zhang, Baochang",
  journal   = "Biomed. Signal Process. Control",
  publisher = "Elsevier BV",
  volume    =  75,
  number    =  103565,
  pages     = "103565",
  month_fake     =  may,
  year      =  2022,
  language  = "en"
}

@ARTICLE{Abuhmed2021-uy,
  title     = "Robust hybrid deep learning models for Alzheimer's progression
               detection",
  author    = "Abuhmed, Tamer and El-Sappagh, Shaker and Alonso, Jose M",
  journal   = "Knowl. Based Syst.",
  publisher = "Elsevier BV",
  volume    =  213,
  number    =  106688,
  pages     = "106688",
  month_fake     =  feb,
  year      =  2021,
  language  = "en"
}

@article{yu2023spatio,
  title={Spatio-temporal graph learning for epidemic prediction},
  author={Yu, Shuo and Xia, Feng and Li, Shihao and Hou, Mingliang and Sheng, Quan Z},
  journal={ACM Transactions on Intelligent Systems and Technology},
  volume={14},
  number={2},
  pages={1--25},
  year={2023},
  publisher={ACM New York, NY}
}
@inproceedings{yu2022graph,
  title={Graph augmentation learning},
  author={Yu, Shuo and Huang, Huafei and Dao, Minh N and Xia, Feng},
  booktitle={Companion Proceedings of the Web Conference 2022},
  pages={1063--1072},
  year={2022}
}

@article{febrinanto2023graph,
  title={Graph lifelong learning: A survey},
  author={Febrinanto, Falih Gozi and Xia, Feng and Moore, Kristen and Thapa, Chandra and Aggarwal, Charu},
  journal={IEEE Computational Intelligence Magazine},
  volume={18},
  number={1},
  pages={32--51},
  year={2023},
  publisher={IEEE}
}

@article{xu2022,
  author={Xu, Chengpei and Jia, Wenjing and Cui, Tingcheng and Wang, Ruomei and Zhang, Yuan-fang and He, Xiangjian},
  journal={IEEE Transactions on Multimedia}, 
  title={Arbitrary-Shape Scene Text Detection via Visual-Relational Rectification and Contour Approximation}, 
  year={2023},
  volume={25},
  number={},
  pages={4052-4066},
  keywords={Feature extraction;Cognition;Visualization;Couplings;Shape;Proposals;Electronic mail;Arbitrary-shape scene text detection;bottom-up method;false positive/negative suppression;relational reasoning},
  doi={10.1109/TMM.2022.3171085}
}

@ARTICLE{pan2024decgan,
  author={Pan, Junren and Zuo, Qiankun and Wang, Bingchuan and Chen, C.L. Philip and Lei, Baiying and Wang, Shuqiang},
  journal={IEEE Transactions on Artificial Intelligence}, 
  title={DecGAN: Decoupling Generative Adversarial Network for Detecting Abnormal Neural Circuits in Alzheimer’s Disease}, 
  year={2024},
  volume={},
  number={},
  pages={1-14},
  keywords={Neural circuits;Integrated circuit modeling;Brain modeling;Feature extraction;Alzheimer's disease;Neuroimaging;Diffusion tensor imaging;Brain Networks;Multimodal Neuroimaging;Hypergraph;Decoupling Algorithm;Sparse Capacity Loss}
}

@article{meng2022multi,
  title={Multi-modal neuroimaging neural network-based feature detection for diagnosis of Alzheimer’s disease},
  author={Meng, Xianglian and Liu, Junlong and Fan, Xiang and Bian, Chenyuan and Wei, Qingpeng and Wang, Ziwei and Liu, Wenjie and Jiao, Zhuqing},
  journal={Frontiers in Aging Neuroscience},
  volume={14},
  pages={911220},
  year={2022},
  publisher={Frontiers Media SA}
}

@article{meng2023research,
  title={Research on early diagnosis of Alzheimer's disease based on dual fusion cluster graph convolutional network},
  author={Meng, Lu and Zhang, Qianqian},
  journal={Biomedical Signal Processing and Control},
  volume={86},
  pages={105212},
  year={2023},
  publisher={Elsevier}
}

@article{yang2023mapping,
  title={Mapping multi-modal brain connectome for brain disorder diagnosis via cross-modal mutual learning},
  author={Yang, Yanwu and Ye, Chenfei and Guo, Xutao and Wu, Tao and Xiang, Yang and Ma, Ting},
  journal={IEEE Transactions on Medical Imaging},
  year={2023},
  publisher={IEEE}
}

@article{zuo2024prior,
  title={Prior-guided adversarial learning with hypergraph for predicting abnormal connections in Alzheimer’s disease},
  author={Zuo, Qiankun and Wu, Huisi and Chen, CL Philip and Lei, Baiying and Wang, Shuqiang},
  journal={IEEE Transactions on Cybernetics},
  year={2024},
  publisher={IEEE}
}
