## Supplementary figures and images for "Multimodal Hyperbolic Graph Learning for Alzheimer’s Disease Detection"

### fig04.png

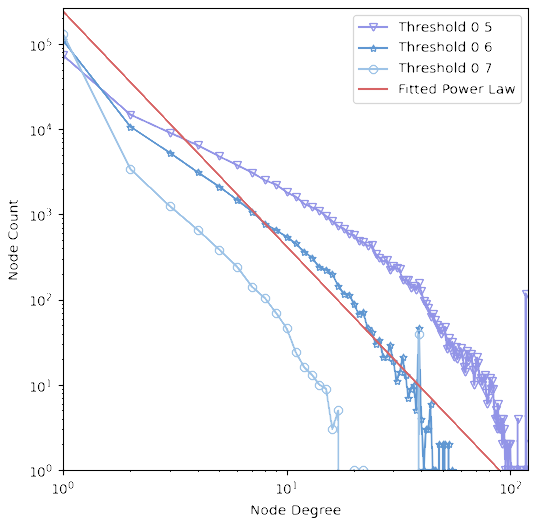

### fig07.png

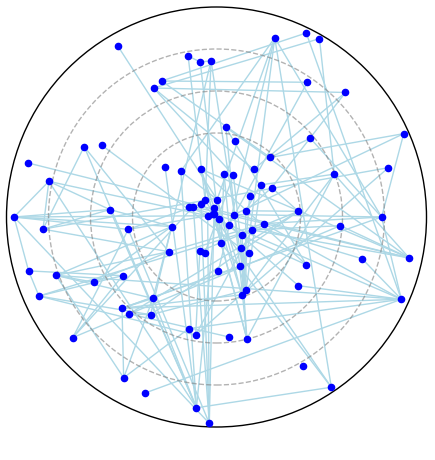

### fig08.png

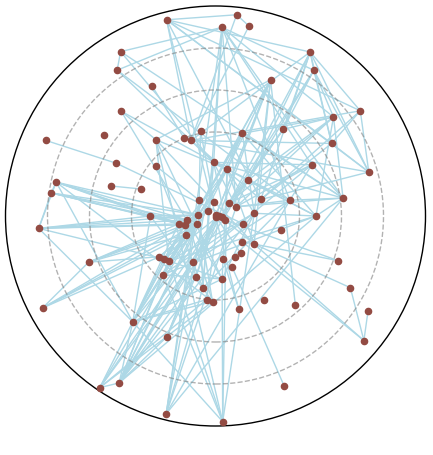

### framework.png

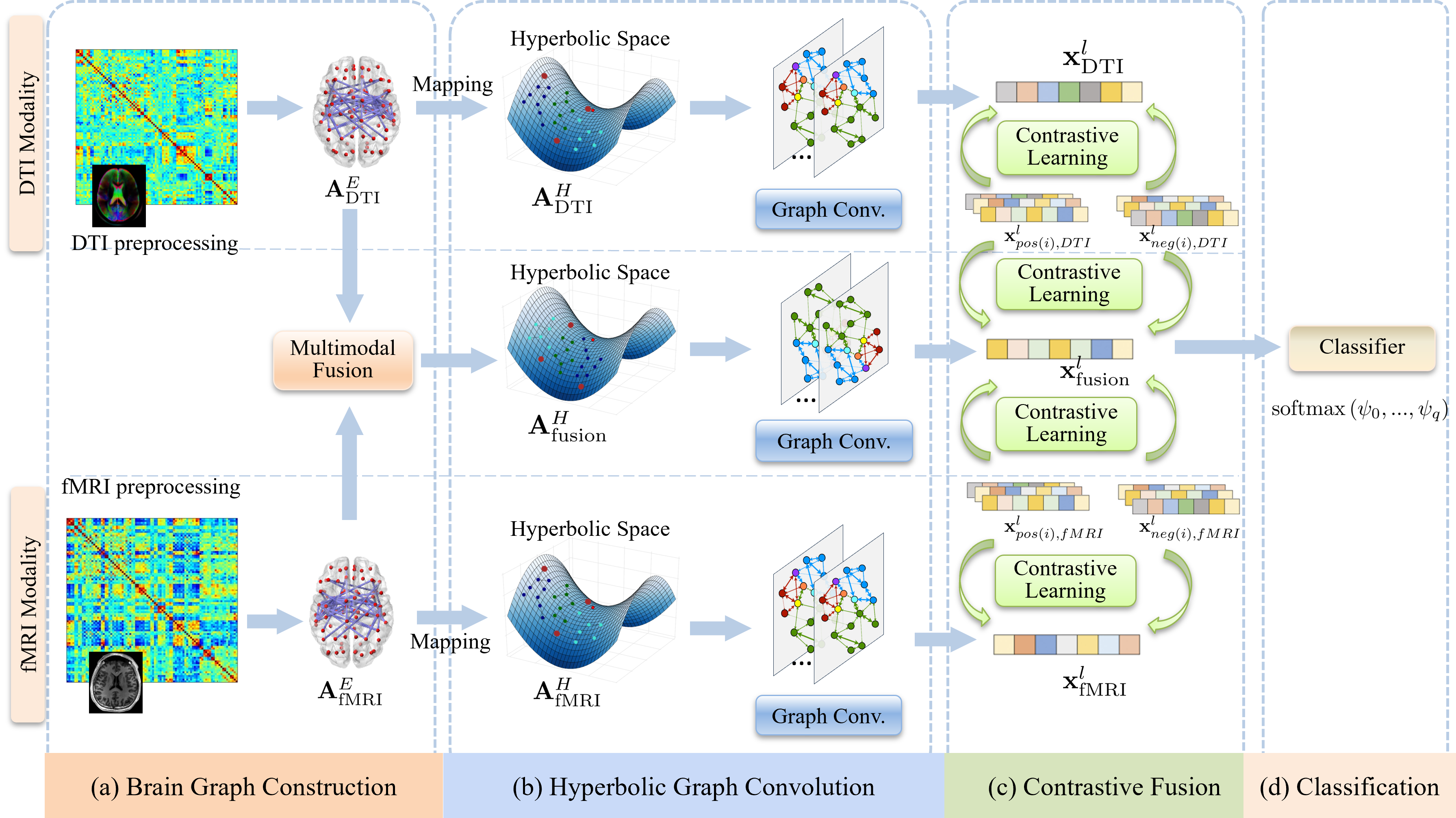

### scale_free.png

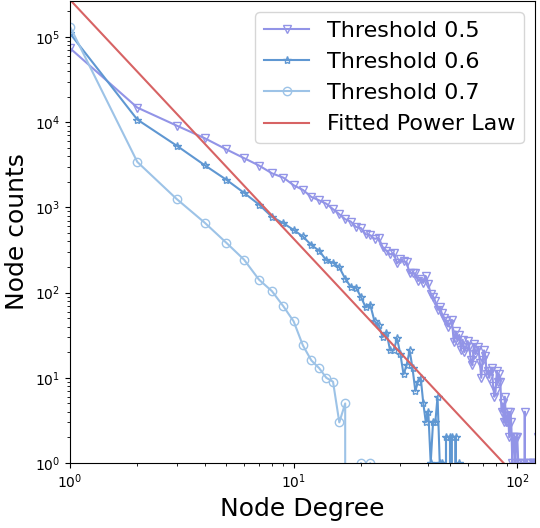
